# COVID-19 convalescents exhibit deficient humoral and T cell responses to variant of concern Spike antigens at 12 month post-infection

**DOI:** 10.1101/2021.11.08.21266035

**Authors:** Pablo Garcia-Valtanen, Christopher M. Hope, Makutiro G. Masavuli, Arthur Eng Lip Yeow, Harikrishnan Balachandran, Zelalem A. Mekonnen, Zahraa Al-Delfi, Arunasingam Abayasingam, David Agapiou, Alberto Ospina Stella, Anupriya Aggarwal, Jason Gummow, Catherine Ferguson, Stephanie O’Connor, Erin M. McCartney, David J. Lynn, Guy Maddern, Eric J Gowans, Benjamin AJ Reddi, David Shaw, Chuan Kok-Lim, Stuart G Turville, Michael R Beard, Daniela Weiskopf, Alessandro Sette, Rowena A. Bull, Simon C. Barry, Branka Grubor-Bauk

## Abstract

**Background:** The duration and magnitude of SARS-CoV-2 immunity after infection, especially with regard to the emergence of new variants of concern (VoC), remains unclear. Here, immune memory to primary infection and immunity to VoC was assessed in mild-COVID-19 convalescents one year after infection and in the absence of viral re-exposure or COVID-19 vaccination.

**Methods:** Serum and PBMC were collected from mild-COVID-19 convalescents at ∼6 and 12 months after a COVID-19 positive PCR (n=43) and from healthy SARS-CoV-2-seronegative controls (n=15-40). Serum titers of RBD and Spike-specific Ig were quantified by ELISA. Virus neutralisation was assessed against homologous, pseudotyped virus and homologous and VoC live viruses. Frequencies of Spike and RBD-specific memory B cells were quantified by flow cytometry. Magnitude of memory T cell responses was quantified and phenotyped by activation-induced marker assay, while T cell functionality was assessed by intracellular cytokine staining using peptides specific to homologous Spike virus antigen and four VoC Spike antigens.

**Findings:** At 12 months after mild-COVID-19, >90% of convalescents remained seropositive for RBD-IgG and 88.9% had circulating RBD-specific memory B cells. Despite this, only 51.2% convalescents had serum neutralising activity against homologous live-SARS-CoV-2 virus, which decreased to 44.2% when tested against live B.1.1.7, 4.6% against B.1.351, 11.6% against P.1 and 16.2%, against B.1.617.2 VoC. Spike and non-Spike-specific T cells were detected in >50% of convalescents with frequency values higher for Spike antigen (95% CI, 0.29-0.68% in CD4^+^ and 0.11-0.35% in CD8^+^ T cells), compared to non-Spike antigens. Despite the high prevalence and maintenance of Spike-specific T cells in Spike ‘high-responder’ convalescents at 12 months, T cell functionality, measured by cytokine expression after stimulation with Spike epitopes corresponding to VoC was severely affected.

**Interpretations:** SARS-CoV-2 immunity is retained in a significant proportion of mild COVID-19 convalescents 12 months post-infection in the absence of re-exposure to the virus. Despite this, changes in the amino acid sequence of the Spike antigen that are present in current VoC result in virus evasion of neutralising antibodies, as well as evasion of functional T cell responses.

**Funding:** This work was funded by project grants from The Hospital Research Foundation and Women’s and Children’s Hospital Foundation, Adelaide, Australia. MGM is THRF Early Career Fellow. BGB is THRF Mid-Career Fellow. This project has been supported partly with Federal funds from the National Institute of Allergy and Infectious Diseases, National Institutes of Health, Department of Health and Human Services, under Contract No. 75N93021C00016 to A.S. and Contract No. 75N9301900065 to A.S, D.W.

**Evidence before this study:** We regularly searched on PubMed and Google Scholar in June-October 2021 using individual or combinations of the terms “long-term immunity”, “SARS-CoV-2”, “antigenic breadth”, “variant of concern” and “COVID-19”. We found studies that had assessed immune correlates at multipe time points after COVID-19 disease onset in convalescents, but not the antigenic breadth of T cells and antibodies and not in relation to VoC. Other immune studies in virus naive vaccinees, or vaccinated convalescents evaluated VoC-specific immunity, but not in convalescents that have not been vaccinated. In summary, we could not find long-term studies providing and in-depth evaluation of functionality of humoral and cell-mediated immunity, combined with addressing the adaptability of these immune players to VoC.

**Added value of this study:** The window of opportunity to conduct studies in COVID-19 convalescents (i.e. natural immunity to SARS-CoV-2) is closing due to mass vaccination programs. Here, in a cohort of unvaccinated mild-COVID-19 convalescents, we conducted a comprehensive, longitudinal, long-term immune study, which included functional assays to assess immune fitness against antigenically different VoC. Importantly, the cohort resided in a SARS-CoV-2-free community for the duration of the study with no subsequent re-exposure or infection. Our findings reveal a deeply weakened humoral response and functional vulnerability of T cell responses to VoC Spike antigens.

**Implications of all the available evidence:** This study provides a valuable snapshot of the quality of SARS-CoV-2 natural immunity and its durability in the context of a pandemic in which new variants continuously emerge and challenge pre-existing immune responses in convalescents and vacinees. Our results serve as a warning that delays in vaccination programs could lead to an increase in re-infection rates of COVID-19 convalescents, caused by virus variants that escape humoral and cell-mediated immune responses. Furthermore, they reinforce the potential benefit of booster vaccination that is tuned to the active variants.

## Introduction

Novel severe acute respiratory syndrome coronavirus 2 (SARS-CoV-2) has infected millions world-wide, causing respiratory coronavirus disease 2019 (COVID-19) and a global pandemic not seen in more than 100 years.^1^ Rapid development and deployment of different COVID-19 vaccines and non-pharmaceutical interventions, such as hard and soft lockdowns are rapidly curbing numbers of daily new infections, hospitalisations and deaths in countries where these measures are implemented.^2-7^ However, while vaccines represent the most likely way out of the pandemic, antibody responses and neutralising activity wane over the months following SARS-CoV-2 primary infection^8,9^ as well as after immunization with current COVID-19 vaccines.^10,11^ SARS-CoV-2 variants with mutations in the Spike protein which enable escape from host antibody responses, add to this problem in convalescents and vaccinees,^12-19^ and have become a major obstacle to ending this pandemic. So far, four variants namely; B.1.1.7 (also known as alpha or UK variant), B.1.351 (beta, RSA), P.1 (gamma, BRA) and B.1.617.2 (delta, IND) have stood out for their ability to spread rapidly across different regions of the world [https://covariants.org/], hence earning them the denomination variant of concern (VoC).

After primary infection and in parallel with the antibody response, symptomatic COVID-19 convalescents generate a robust CD4^+^ and CD8^+^ memory T cell response which targets a wider range of antigens and epitopes than that covered by antibodies.^20-24^ Importantly, the breath of SARS-CoV-2-specific T cell epitopes appears to be less sensitive to mutations present in VoC.^25,26^ It is unclear to what extent T cells can protect from re-infection and progression to severe COVID-19. However, it is likely that T cell responses in convalescents, which target most SARS-CoV-2 antigens,^20^ could afford some of level of protection for many months, even years. In fact, SARS-CoV-specific T cells can be detected in convalescents for almost two decades.^27^

While current vaccines are highly effective in preventing severe disease and death, and booster vaccinations may temporarily circumvent dwindling efficacy over time,^28^ next generation vaccines, that can prevent virus transmission, are likely needed to end the pandemic.^29,30^ Long-term studies of the evolution of immune correlates in COVID-19 convalescents, where the immune system has encountered an active live virus infection in the presence of all its antigens, are necessary to elucidate the fine specificities and immune functionality of antibody and T cell responses. In particular, the adaptability of pre-existing immunity to mutated Spike antigens, present in VoC, is the key piece of information that is still unanswered.

Compared to the majority of the world, South Australia is in a unique position to undertake studies on mid- to long-term immunity of COVID-19 due to 1) early and strict border control measures with other countries, and other states within Australia, which were enforced by health authorities in 2020-21, thus eliminating local transmission of the virus in the community, and 2) South Australia has maintained a high testing rate with a total test count of >2.2 M with only 899 positive cases of which only 9 were caused by unknown, locally-acquired contacts (accessed on 23/09/2021).^31^

We present a COVID-19 immunity study at 12 months after PCR confirmed SARS-CoV-2 infection and in the complete absence of community transmission in a South Australian cohort of 43 mild COVID-19 convalescents. An in-depth evaluation of multi-isotype antibody responses, homologous pseudotyped virus, homologous and VoC live-virus serum neutralisation activity, RBD-specific B cell populations and Spike and non-Spike SARS-CoV-2 specific CD8^+^ and CD4^+^ T cell immunity against ancestral and VoC antigenic epitopes, was undertaken. Results were compared to age- and gender-matched COVID-19 naïve, healthy individuals and to COVID-19 convalescent responses at 6 months after infection in the same cohort.

## Methods

### Study design and setting

Individuals who tested positive in a nasopharyngeal swab SARS-CoV-2 RT-qPCR test in March-April 2020 at South Australian Pathology Services (Adelaide, Australia) were asked to participate in a longitudinal study to assess SARS-CoV-2-specific humoral and immune cell immune correlates (figure 1A). In this study, whole blood specimens from 43 participants (19 male, 24 female) of (95% CI, 44.8-55.4) years of age (figure S1) who presented mild COVID-19 symptoms according to NIH guidelines (https://www.covid19treatmentguidelines.nih.gov/overview/clinical-spectrum/) were sampled and processed at two time points 6 months apart after PCR positive test. All samples were coded and de-identified for analysis. None of the participants in this study tested positive for COVID-19 after their initial positive test. Age- and gender-matched seronegative (SARS-CoV-2 RBD and Spike) healthy controls (n=15) were included to provide baseline levels of different immune correlates (figure S1). Prior to this, a portion of our mild-COVID-19 convalescents participated in a separate study that included serological, immunophenotyping and whole blood RNAseq analysis.^32^

**Figure 1.**
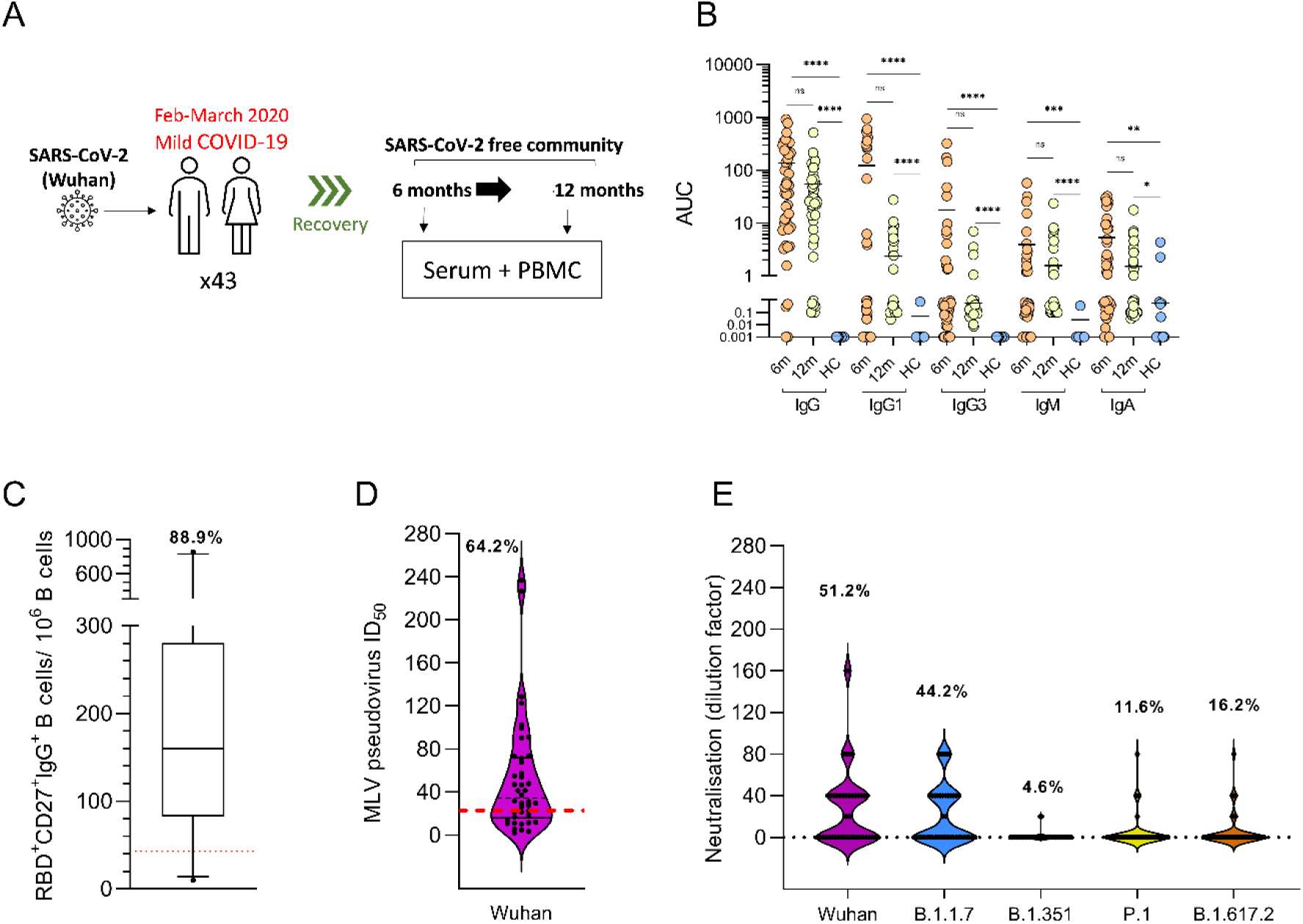
Circulating RBD-specific antibodies, memory B cells frequency and serum SARS-CoV-2 neutralisation activity at 12 months after PCR positive test. **(A)** Forty-three SARS-CoV-2 infected individuals who presented mild-COVID-19 symptoms were recruited after a PCR positive test and serum and PBMCs were sampled at 6 and 12 months. **(B)** Serum RBD-specific antibody titers, per Ig isotype, reported as area under the curve (AUC) units. Circles represent AUC individual patient values (n=43 at 6 months, orange, and 12 months, yellow, n=15 for healthy controls, blue), with mean value denoted by a horizontal black line. Seronegative samples were assigned a value of 0.001 data visualisation purposes. **(C)** SARS-CoV-2 RBD-specific (n=28) memory B cells (CD27^+^) were quantified 12 month post-infectoion with corresponding specific tetramers and further characterised as IgG^+^. Cell population-specific background was calculated with healthy control PBMCs and shown as a red dashed line.^36^ **(D)** Serum neutralisition ID_50_ of SARS-CoV-2, MLV pseudovirus particles expressing infectious homologous Spike sequence (Wuhan) in mild-COVID-19 convalescent sera (n=42) at 12 months after positive COVID-19 test. Positive neutralization percentage (indicated above figure) activity cut-off (ID_50_= 22.61) was calculated from 19 healthy control samples, shown as a red dashed line. **(E)** Patient serum neutralisation end-point cut-off titers (highest dilution factor that yields ≥50% inhibition of cell death after live virus infection) at 12 months against Wuhan, B.1.1.7, B.1.351, P.1 and B.1.617.2 live virus particles. Forty was the initial dilution for all serum samples. Neutralisation activity was considered negative, value of zero, when neutralisation of initial serum dilution was <50%. ^*, **, ***^ and ^****^ denote *P* values < 0.05, 0.01, 0.001, 0.0001 respectively. ns = not significant.

Blood collection and processing at the Royal Adelaide Hospital, Women’s and Children’s Hospital, and The University of Adelaide were performed as previously described.^33^ Blood was collected via phlebotomy in serum separator tubes (no additives) or ethylenediaminetetraacetic acid (EDTA) tubes and processed for serum, plasma and peripheral blood mononuclear cells (PBMCs) isolation.

Study protocols were approved by the Central Adelaide Clinical Human Research Ethics Committee (#13050) and the Women’s and Children’s Health Network Human research ethics (protocol HREC/19/WCHN/65), Adelaide, Australia. All participants provided written informed consent in accordance with the Declaration of Helsinki and procedures were carried out following the approved guidelines.

### SARS-CoV-2 RBD and Spike protein production

Prefusion SARS-CoV-2 Spike ectodomain (isolate WHU1, residues 1-1208) with HexaPro mutations (kindly provided by Dr Adam Wheatley)^34^ was used in ELISA. SARS-CoV-2 Spike transmembrane domain removed and a C-terminal His-tag (residues 1-1273) (kindly provided by Prof Florian Krammer)^35^ was used for flow cytometric detection of Spike-specific B cells. SARS-CoV-2 RBD with C-terminal His-tag (residues 319-541; kindly provided by Prof Florian Krammer)^35^ was used in ELISA and flow cytometry. Recombinant proteins were overexpressed in Expi293 cells (Thermo Fisher) and 72 hrs later purified by Ni-NTA affinity and size-exclusion chromatography.^19^ Purified proteins were quantified using the Bradford protein assay (Bio-Rad) and analysed by SDS-PAGE and Western blot before being stored at - 80°C. Recombinant RBD was biotinylated using the Avitag as described by the manufacturer (Genecopeia), while the Spike protein was biotinylated using an EZ-Link™ Sulfo-NHS-Biotin kit.

### SARS-CoV-2 RBD ELISA

To normalise assay variance between individual experiments, standardized positive and negative controls were used. The threshold was set from sera collected from 40 healthy individuals prior to 2019 (mean + 2SD), while the positive control was sera from COVID-19 convalescents, collected 12 weeks after positive COVID-19 PCR test. MaxiSorp 96-well plates were coated overnight at 4°C with 5 μg/mL of recombinant RBD protein and blocked with 5% w/v skim milk in 0.05% Tween-20/PBS (PBST) at room temperature. Heat inactivated patient sera were serially diluted in blocking buffer, added and incubated for 2h at room temperature, followed by four washes in 0.05% PBST. Secondary antibodies were diluted in 5% skim milk in PBST as follows: Goat anti-Human IgG (H+L) Secondary Antibody, HRP (1:30,000; Invitrogen); Mouse Anti-Human IgG1 Fc-HRP (1:5000, Southern Biotech), Mouse Anti-Human IgG3 Hinge-HRP (1:5,000; Southern Biotech); Goat anti-human IgM HRP (1:5,000; Sigma): Goat anti-human IgA HRP antibody (1:5,000; Sigma) and incubated for 1 hour at room temperature, followed by four washes with PBS-T. Plates were developed with 1-Step™ Ultra TMB Substrate (ThermoFisher Scientific) and the reaction stopped with 2M sulphuric acid. Absorbance was measured at 450nm using Synergy HTX Multi-Mode Microplate Reader. SARS-CoV-2 Spike and RBD endpoint titers were calculated and expressed as area under the curve (AUC) using the mean optical density (OD) reading of the first dilution from healthy normal human sera as the baseline cut-off for seropositivity. AUC calculations were performed using Prism GraphPad.

### Detection of Spike- and RBD-specific memory B cells

The staining method used for the detection of RBD and Spike memory B cells has been described previously.^36^ In brief, biotinylated RBD and Spike proteins were incubated with Streptavidin-PE (SA-PE; Molecular probes; ThermoFisher Scientific) and Streptavidin-APC (SA-APC; BD Pharmingen) in a molar ratio of 4:1 and 2:1, respectively. Cryopreserved PBMCs were thawed rapidly in a 37°C water bath and washed with pre-warmed RPMI media supplemented with 2 mM L-glutamine, 50 IU/mL penicillin, 50 μg/mL streptomycin, and 10% heat-inactivated fetal calf serum (Sigma) then washed twice. A maximum of 1 × 10^7^ cells were stained with Fixable Viability Stain 700 (FVS700) (BD Bioscience in a 1:1000 dilution), 5 µL Human Fc block (BD Bioscience) per 2 × 10^6^ cells, 1 µg/mL each of RBD and Spike tetramers, 5 µL each of CD21 BV421, IgD BV510, CD10 BV605, CD19 BV711 and CD20 APC-H7, 8 µL of IgG BV786, 2 µL each of CD27 PE-CF594 and CD38 PE-Cy7, 2.5 µL HLA-DR BB515 and 0.5 µL CD3 BB700 (BD Bioscience). Cells were washed, resuspended in FACS wash buffer and the data acquired on BD FACSAria™ III. Data analysis was performed using FlowJo version 10.7.1 (TreeStar).

### SARS-CoV-2 pseudovirus neutralisation assay

Pseudovirus neutralisation assays were performed as previously described.^36^ In brief, SARS-CoV-2 pseudo-particles were generated by co-transfecting expression plasmids containing SARS-CoV-2 Spike (kindly provided by Dr Markus Hoffmann)^37^ and the MLV gag/pol and luciferase vectors (kindly provided by Prof. Francois-Loic Cosset)^38,39^ in CD81KO 293T cells (kindly provided by Dr Joe Grove),^40^ using mammalian Calphos transfection kit (Takara Bio). Transfected cells were then incubated at 32°C, 5% CO_2_ with the culture supernatants containing SARS-CoV-2 virus pseudoparticles (SARS-2pp) and harvested 48 hours post-transfection, concentrated 10-fold using 100,000 MWCO Vivaspin centrifugal concentrators (Sartorius) and stored at –80°C.

For neutralisation assays, SARS-2pp were diluted 50-fold and incubated for 1hr with heat-inactivated patient serum, followed by the addition of polybrene at a final concentration of 4µg/mL (Sigma-Aldrich), prior to addition to 293T-ACE2 over-expressing cells (kindly provided by A/Prof Jesse Bloom).^41^ 293T-ACE2 cells were seeded 24 h earlier at 1.5 × 10^4^ cells per well in 96-well white flat-bottom plates (Sigma-Aldrich). Cells were spinoculated at 800 g for two hours and incubated for two hours at 37°C, prior to media change. After 72 h, cells were lysed with a lysis buffer (Promega) and Bright Glo reagent (Promega) was added at a 1:1 ratio. Luminescence (RLU) was measured using CLARIOstar microplate reader (BMG Labtech). Neutralisation assays were performed in triplicate and outliers were excluded using the modified z-score method.^42^ Percentage neutralisation of SARS-2pp was calculated as (1 – RLU treatment/RLU no treatment) × 100 and the half-maxium inhibitory serum dilution (ID_50_) was calculated for each sample. An ID_50_ positive neutralisation cut-off of 22.61 was determined using ID_50_values obtained from 19 unexposed healthy participants (mean + 2 SD).^36^ ID_50_ for serum was calculated using a non-linear regression model (GraphPad Prism).

### SARS-CoV-2 live-virus neutralisation assay

HEK-ACE2/TMPRSS cells (Clone 24)^43^ were seeded in 384-well plates at 5 × 10^3^ cells/well in the presence of the live cell nuclear stain Hoechst-33342 dye (NucBlue, Invitrogen) at a concentration of 5% v/v. Two-fold dilutions of patient plasma samples were mixed with an equal volume of SARS-CoV-2 virus solution (1.25 × 10^4^ TCID50/ml) and incubated at 37°C for 1 hour before adding 40 μl, in duplicate, to the cells (final MOI = 0.05). Viral variants used included the key variants of concern; Alpha (B.1.1.7), Beta (B.1.351), Gamma (P1) and Delta (B.1.617.2), as well as ‘wild-type’ control virus (A.2.2) from clade A and presenting no aa mutations in Spike (similar to Wuhan ancestral variant). Plates were incubated for 24 hours post infection and entire wells were imaged by high-content fluorescence microscopy, cell counts obtained with automated image analysis software, and the percentage of virus neutralisation was calculated with the formula: %N = (D-(1-Q)) × 100/D, as previously described.^43^ An average %N > 50% was defined as having neutralising activity.

### SARS-CoV-2 and CMV peptide megapools

SARS-CoV-2 and CMV peptide megapools were kindly provided by Prof Alessandro Sette (La Jolla Institute of Immunology, CA, USA).^23^ For SARS-CoV-2 whole proteome, CD8-specific peptide pools, 628 peptides restricted to the 12 most common HLA-A and HLA-B alleles and partially covering the sequences of nsp1, nsp2, PLpro, nsp4, nsp6, nsp7, nucleocapsid phosphoprotein, 3CL, nsp8, nsp9, nsp10, nsp14, RdRpol, Hel, nsp15, nsp16, surface glycoprotein, ORF3a, ORF10, ORF6, ORF7a, ORF8, envelope protein, and membrane glycoprotein were predicted *in silico* as previously described.^21^ Peptides were divided into two separate megapools, CD8_A and CD8_B. In Spike peptide pool, 15-mer peptides overlapping by 10 amino acids and covering the entire Spike protein sequence were used (total of 253 peptides). For the non-Spike SARS-CoV-2 CD4 megapool 221 15-mer restricted to seven common HLA-DR (Class II HLA) alleles and covering the entire SARS-CoV-2 proteome, except for the Spike protein, were predicted *in silico* as described previously.^21^ The CD8 and CD4 human Cytomegalovirus (CMV) megapools used as a virus-positive control response consist of HLA-restricted 204 and 141 peptides.^44^ All peptides were synthesized and resuspended in DMSO at 1 mg/ml.

### Activation induced cell marker (AIM) T cell assay

Thawed PBMCs were rested for 2 hours at 370°C, 5% CO_2_ in complete RPMI (cRPMI) medium (40 U/mL penicillin, 40 ug/mL streptomycin, 2 mM L-Glutamine) with 5% (v/v) heat-inactivated human AB serum. Cells were then plated at 10^6^ PBMC/well in u-bottom 96-well plates and stimulated with 1 µg/mL of different SARS-CoV-2-megapools. Combined CD4 and CD8 cytomegalovirus (CMV) megapool (1 µg/mL) and PHA 10µg/mL (Sigma Aldrich), were included as positive controls. An equimolar amount of dimethyl sulfoxide (DMSO, vehicle) was used as a negative control. PBMC were stimulated for 24 h at 37^°^C, 5% CO_2_, washed and stained with Zombie Green Fixable Live/Dead Stain (L/D,Biolegend) for 20 min, RT, in the dark. Cells were then washed and stained with (CD3 BUV737, CD4 BUV496, CD8 BUV395, CD14 FITC, CD19 FITC, CD45RA BV650, CCR7 (CD197) APC, CD69 PE, CD134 (OX40) PE-Cy7, CD137 (41-BB) BV421) for 20min, at room temperature in the dark. Fluorescence minus one (FMO) control for antigens: CD45RA, CCR7, CD134, CD69 and CD137 were added to PHA stimulated cells. PBMC were washed and FACS Fix (0.4%PFA, 20g/L Glucose, Sodium Azide 0.02% in PBS) was added for 20 min at room temperature, in the dark. Fixed cells were washed, resuspended in FACS wash buffer and data was acquired on BD FACS Symphony. Data analysis was performed using FCS Express ™ (DeNovo Software, Pasadena, CA, USA). All percentages of activated cells were calculated subtracting unspecfic DMSO background for each cell phenotype and individual patient.

### Variant of concern Spike-specific peptide pools

SARS-CoV-2 Spike PepTivator® protein pools (Miltenyi Biotec, Gladbach, GER) were utilised to test immune reactivity of COVID-19 convalescent T cells to mutated Spike epitopes present in four VoC.^26,45^ All peptide pools consisted of 15-mer peptides with 11 aa overlap covering Spike protein sequences affected by mutations in each VoC. Four VoC peptide pools corresponding to mutated Spike sequences in SARS-CoV-2 variants B.1.1.7, B.1.351, P.1 and B.1.617.2 (34, 30, 41 and 32 peptides, respectively) and four corresponding control/reference pools with Wuhan aa sequences were compared in parallel along with whole Spike peptide pool as positive control (pool described above). Mutations and deletions represented in mutated pools are summarized in Table S1. For the assays, lyophilised peptides were resuspended in sterile miliQ water as per the manufacturer’s instructions at 30 nM (50 µg/mL), aliquoted and stored at -80°C until used.

### Spike-specific T follicular helper cell quantification and intracellular cytokine staining in Spike high responder convalescents

According to AIM assay results, convalescents with Spike-specific CD4^+^ and CD8^+^ T cell frequencies above the mean plus 3^*^standard deviations in the healthy control group were identified. Double responders (meeting criterion for both CD4^+^ and CD8^+^ T cells) and high CD4^+^ with available PBMC samples were selected (n=15) for further analysis. Following methods similar to other published prior to this study,^46^ PBMCs were thawed and prepared for cell culture as described for the AIM assay. Cells were pre-treated with 0.555 µg/mL of anti-CD40 blocking antibody (HB14, Miltenyi Biotec) for 15 min. Then, peptide pools were added to a final concentration of 1 µg/mL (making anti-CD40 concentration 0.5µg/mL for the remainder of the stimulation period). After a 24 hr incubation 2 µM GolgiStop™ (containing monensin, BD, 554724) and 1µg/mL GolgiPlug™ (containing Brefeldin A, BD, 555029) were added to the cells and incubated for an additional 4 hrs. Cells were then co-incubated with Fixable Viability stain 780 (BD) and Fc Block (BD) for 20 min, RT, in the dark, washed with FACS buffer solution and stained with surface stain mix (CD3 BUV737, CD4 BUV496, CD8 BUV395, CXCR5 BUV563, CD14 APC-Cy7, CD20 APC-Cy7) for 20 min, RT, in the dark. Cells were washed with PBS and subsequently fixed and permeabilized with Cytofix/Cytoperm™ (BD, 51-2090KZ) for 20 min, RT, in the dark. Cells were then washed with Perm/Wash™ (BD, 51-2091KZ) and stained with ICS stain Mix (CD154 PE, IFNγ PE-Cy7, TNFα APC, PRF1 FITC, IL-2 BV711, GZMB BV421) for 20 min, RT, in the dark. Cells were then washed twice with Perm/Wash™ and once with PBS. Finally, cells were resuspended in PBS and kept at 4 °C until data was acquired on BD FACS Symphony. Data analysis was performed using FCS Express ™ (DeNovo Software, Pasadena, CA, USA).

Fold-change values in ICS data for VoC peptide pools and reference pools were calculated as follows: Fold Change (MUT/REF)= VoC pool % of cytokine positive cells/ Reference pool % of cytokine positive cells. From fold-change values between 0 and 1, negative fold-change values were calculated: Negative fold= 1/(Fold change [MUT/REF]).

### Statistics, replicates and sample-size estimation

All mild-COVID-19 patient samples available at the time points reported in the study from the South Australian cohort (n=43) were used in this study and, therefore, no *pre hoc* power calculations were carried out to determine the sample size. A sufficient number of healthy controls (n=15) were recruited to establish meaningful comparisons and assay cut-off and baseline levels consistent with current literature. Additional healthy controls (sampled prior to 2019) were used to validate serological, virus neutralisation and B cell staining assays (see Methods section). All statistical analyses were performed using GraphPad Prism 9.0.0 (San Diego, CA, US). No assumptions were made about the distribution of the data sets; non-parametric tests were used in all cases for comparisons. Accordingly, two-tailed Mann-Whitney’s or Welch’s tests were applied for pair-wise comparisons or Krustal-Wallis test for unpaired comparisons (HC vs patient data) of antibody and T cell frequency data. A non-linear regression model was used to calculate individual patient ID_50_values from corresponding pseudovirus particles neutralisation assay data.^47^ A mixed-effects analysis was performed to carry out all pair-wise comparisons in memory B cell frequency data. Conservative multiple comparisons corrections were not applied in statistical analysis in order to avoid obscuring existing associations between different immune correlates.

### Randomisation

Since all samples that were available in South Australia were used in the study, no randomisation was performed in the experiments.

### Blinding

No experiments were blinded in this study.

## RESULTS

### Longitudinal SARS-CoV-2 humoral responses in mild-COVID-19 convalescents

The receptor binding domain (RBD) of SARS-CoV-2 Spike protein is the main target of neutralising antibodies (nAb), and nAb titers decline in the months after COVID-19 infection. Although not statistically significant, RBD-specific serum immunoglobulin had a downward trend for all isotypes between the time points analysed (figure 1B). The RBD IgG, IgG1 and IgG3 AUC titers decline was (95% CI, from 73.0-200.4 to 27.7-81.9), (from 66.8-218.2 to 0.8-3.8) and (from 0.0-44.3 to 0.18-0.92), respectively (figure 1B). In comparison with healthy seronegative control AUC values, RBD seropositivity for IgG isotypes was present in the majority of COVID-19 convalescents. This trend positively correlated with the presence of above background levels of circulating memory (CD27^+^) B cells expressing RBD-specific surface IgG in 88.9% of individuals in the COVID-19 convalescent cohort (95% CI, 153-336 cells/ 10^6^ B cells) compared to healthy controls (95% CI, 0.0-27.9 cells/10^6^ B cells) (figure 1C), indicative of the existence of long-term SARS-CoV-2-specific humoral immunity 12 months post-infection. Spike-specific IgG^+^ B cells were also elevated in COVID-19 convalescents, but healthy control background frequencies were also higher (figure S2), likely due to cross-reactivity. RBD- and Spike-specific non-IgG^+^ B cell frequencies were present a low rates (figure S2).

Obtaining long-term serum neutralisation data from communities free of circulating SARS-CoV-2, such as in this study, is difficult in other cohorts in the context of this pandemic. In our cohort, sera in 64% of convalescents at 12 months yielded ID_50_ neutralisation titers significantly above healthy background levels against the pseudotyped virus bearing a Wuhan-like Spike protein, which is the same as the prevalent virus present in the community when study participants were infected (figure 1D).

Since early 2020, when the study participants were recruited, four VoC, namely, Alpha (B.1.1.7), Beta (B.1.351), Gamma (P1) and Delta (B.1.617.2), have dominated the landscape of COVID-19 outbreaks worldwide. Neutralisation titers against live-Wuhan virus were 16.8-40.8 (95% CI) with 51.2% of convalescents at 12 months presenting positive neutralising activity (figure 1E). Values were similar to those against pseudotyped virus particles bearing the same Spike protein sequence in figure 1D. As expected the percentage of patients with positive neutralising activity against B.1.1.7 virus did not differ significantly (44.2%) to Wuhan with titers ranging 12.8-29.9 (95% CI). However, a very significant drop in serum neutralisation titers was observed for live virus B.1351 (95% CI, 0.0-2.2), P.1 (0.2-9.9) and B.1.617.2 variants (1.1-10.9) with only 4.6%, 11.6% and 16.2% of convalescents, respectively, exhibiting positive neutralisation activity (figure 1E).

This is indicative that despite a high prevalence of RBD-seropositivity, existence of circulating memory B cells and homologous virus neutralisation activity among COVID19 convalescents, functional humoral responses to VoC are significantly reduced at 12 months post-infection.

### Longitudinal quantification and phenotyping of SARS-CoV-2 T cell responses in mild-COVID-19 convalescents

Along with antibody responses, the majority of COVID-19 convalescents develop SARS-CoV-2 Spike- and non-Spike-specific T cell responses.^48^ These T cells can be detected and quantified by means of activation-induced marker (AIM) assays using SARS-CoV-2 antigen peptide pools and flow cytometric analysis (figure S3).^20,22,23^ For accurate interpretation in SARS-CoV-2 AIM assays, naïve healthy controls must be included in the analysis to establish assay baseline levels that arise from previous immunity to unrelated antigens/pathogens, and particularly to seasonal human coronaviruses.^49^ In our cohort, the relative frequency of CD4^+^ and CD8^+^ T cells did not differ between the two time points (6 and 12 months) or between COVID-19 convalescents and healthy controls (figure S4A).

AIM assays revealed that the frequency of circulating Spike-specific CD4^+^ T cells did not significantly decrease between 6 and 12 moths post-infection (95% CI, from 0.35-0.91 at 5-6 months to 0.29-0.68) (figure 2A). The same trend was observed for non-Spike antigen CD4^+^ T cells (from 0.22-0.43 to 0.18-0.37) and for the combined CD4^+^ T cell response, Spike + non-Spike (from 0.60-1.33 to 0.49-1.04) (figure 2A).

**Figure 2.**
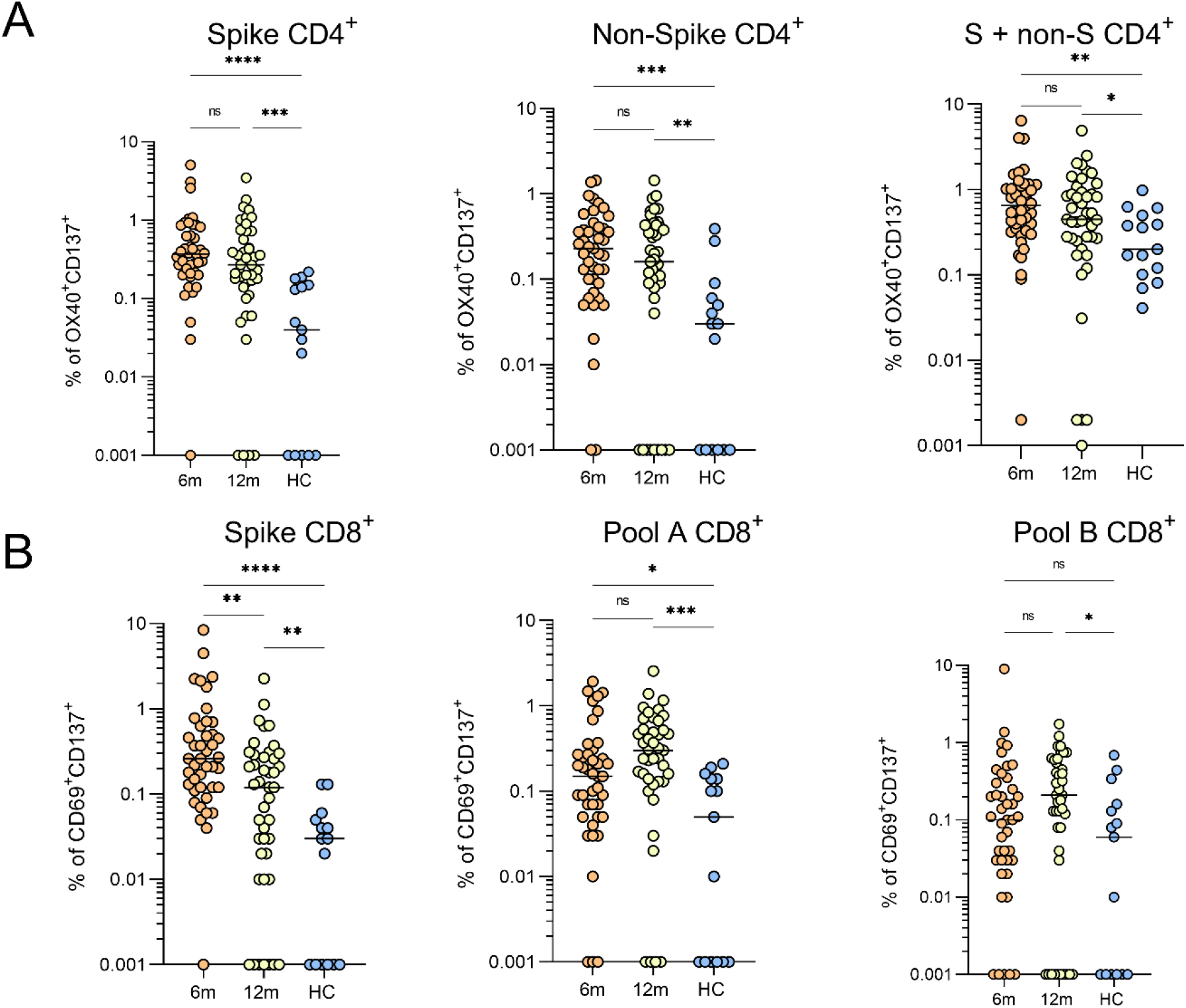
SARS-CoV-2-specific CD4^+^ and CD8^+^ T cell frequencies in mild-COVID-19 convalescents at 6 and 12 months after PCR positive test. (**A**) Percentage of activated CD4^+^ T cells (OX40^+^CD137^+^) after stimulation with Spike, non-Spike and cumulative (Spike + non-Spike) antigen peptide pools within the total CD3^+^CD4^+^ T cell population of PBMCs in individual mild-COVID-19 patients (n=43) at 6 and 12 months after COVID-19 positive test (orange and yellow) and healthy controls (n=15, in blue). (**B**) Percentage of activated CD8^+^ T cells (CD69^+^CD137^+^) after stimulation with Spike, whole proteome, A and B antigen peptide pools within the total CD3^+^CD8^+^ T cell population of PBMCs in same samples as in (A). Dots in a represent patient or healthy control individual values. Averages are denoted by a horizontal line and statistically significant differences between patient and healthy controls indicated by asterisks. ^*, **, ***^ and ^****^ denote *P* values < 0.05, 0.01, 0.001 and 0.0001, respectively. ns = not significant.

Comparatively, the reduction of Spike-specific CD8^+^ T cell frequency over time was more pronounced and statistically significant (95% CI, from 0.28-1.19 to 0.11-0.35, p<0.01) (figure 3B). No statistically significant differences were observed for the frequencies of CD8^+^ T cell reacting to whole SARS-CoV-2 proteome pools A and B (figure 2B). Convalescent AIM results were compared to naïve healthy controls in all instances, corroborating significantly lower T cell frequencies (baseline levels), consistent with previous AIM studies using the same peptide pools.^22,23^ In addition, a positive control consisting of CMV protein peptides^22,23,44^ was included for all healthy controls and 12 month convalescent data samples. As expected, a high percentage of individuals had a relatively high percentage of CMV-specific T cells (both CD4^+^ and CD8^+^) with no significant differences between healthy and convalescent cohorts (figure S4B). These results are in line with other AIM longitudinal COVID-19 studies spanning from acute phase to up to 10 months after disease onset and indicate sustained maintenance of T cell responses one year post-infection.^22,50^

**Figure 3.**
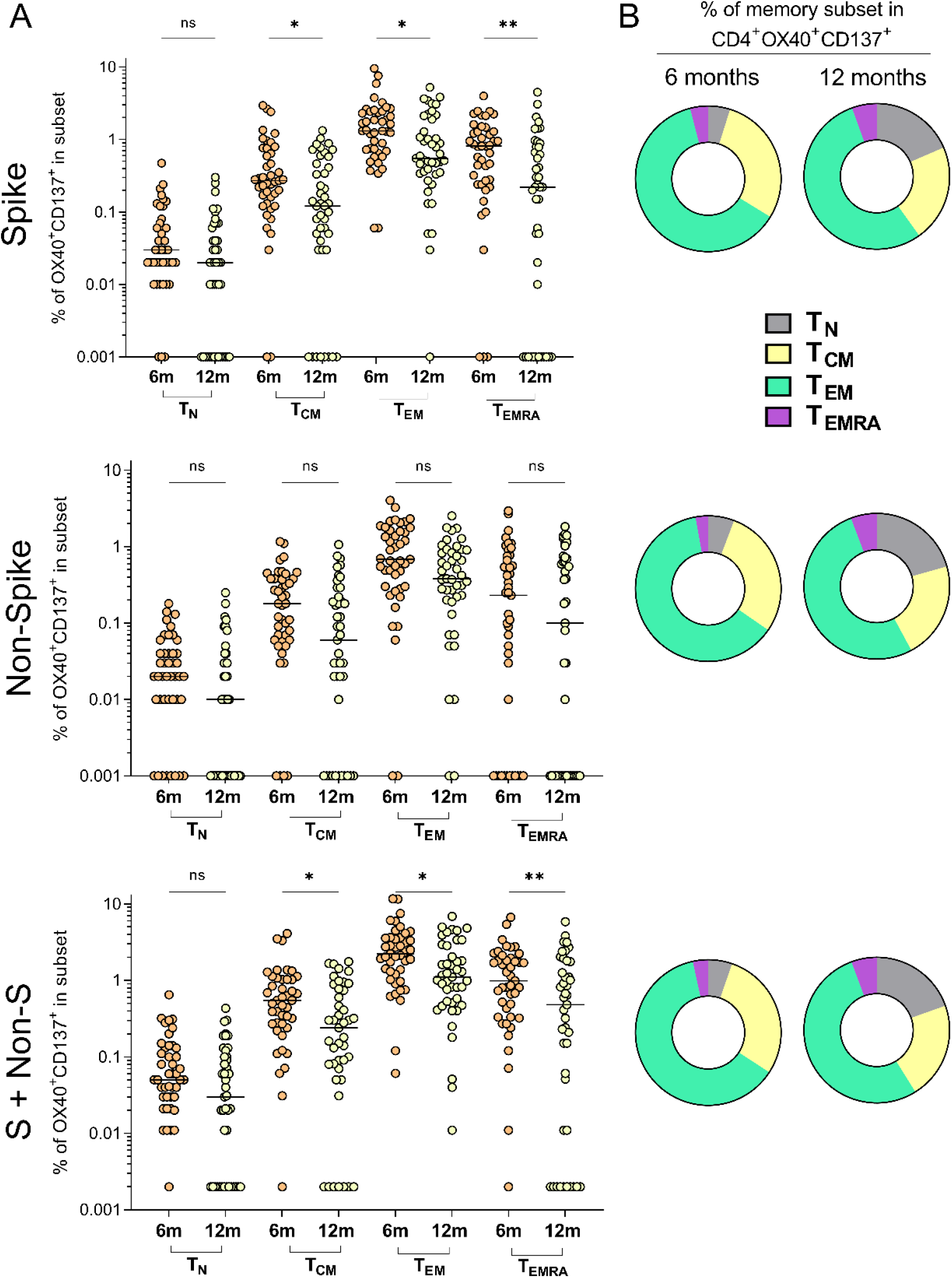
Memory immunophenotyping and quantification of SARS-CoV-2-specific CD4^+^ T cells in mild-COVID-19 convalescents at 6 and 12 months after PCR positive test. **(A)** Memory immunophenotype based on expression of surface CCR7 and CD45RA of SARS-CoV-2 Spike-, non-Spike- and cumulative (Spike + non-Spike)-specific CD4^+^ T cells detected in Figure 2A. Cells were classified as follows: naïve (T_N_, CCR7^+^CD45RA^+^), central memory (T_CM_, CCR7^+^CD45RA^-^), effector memory (T_EM_, CCR7^-^CD45RA^-^) and terminally differentiated effector memory cells re-expressing CD45RA (T_EMRA_, CCR7^-^CD45RA^+^). Frequencies are indicated as percentage of total SARS-CoV-2 antigen-specific CD4^+^ T cells within the total pool of immune cells with same phenotype in the patient’s PBMCs.**(B)** Doughnut charts indicating the proportion (%) of each immune phenotype in (A) of the total of SARS-CoV-2-specific CD4^+^ T cells for each antigen. In (A) circles represent patient individual values. Averages are denoted by a horizontal line and statistically significant differences between time points indicated by asterisks. ^*^ and ^**^denote *P* values < 0.05 and 0.01, respectively. ns = not significant.

Next, we evaluated the memory phenotype of SARS-CoV-2-specific CD4^+^ and CD8^+^ T cells to support stronger conclusions on the specific roles of each population in the maintainance of SARS-CoV-2 immunity. Measuring the expression of surface markers CCR7 and CD45RA allows for the identification of different subsets of memory T cells, with different function, kinetics and frequency depending on the immune status, pathology and age.^51^ In the AIM assay, we assessed the frequencies of SARS-CoV-2-specific T cells within the total pool of central memory (T_CM,_ CCR7^+^CD45RA^-^) T cells, effector memory T cells (T_EM_, CCR7^-^CD45RA^-^), terminally differentiated or terminal effector T cells (T_EMRA_, CCR7^-^CD45RA^+^) and naïve T cells (T_N,_ CCR7^+^CD45RA^+^) in both CD4^+^ and CD8^+^ T pools. Between 6 and 12 months post infection, the frequency of Spike-specific CD4^+^ T cells were significantly reduced in all three memory compartments, T_CM_ (95% CI, from 0.32-0.73 to 0.17-0.37, p<0.05), T_EM_ (1.2-2.4, 0.75-1.52, p<0.05) and T_EMRA_ (0.67-1.20, 0.29-0.85, p<0.01) (figure 3A). Although the trend is similar for non-Spike CD4^+^ T cell memory subsets, the differences between time points were not statistically significant (figure 4A). The combined data (Spike + non-Spike CD4^+^ T cell subsets) showed statistically significant decrease in SARS-CoV-2-specific CD4^+^ T_CM_ (0.51-1.06, 0.28-0.60, p<0.05), T_EM_ (2.11-3.68, 1.22-2.24, p<0.05) and T_EMRA_ (0.98-1.83, 0.58-1.37, p<0.01) (figure 3A). Importantly, effector memory T cell (T_EM_) phenotype was clearly predominant in SARS-CoV-2-specific CD4^+^ T cells making up >60% and >50% of all CD4^+^OX40^+^CD137^+^, at 6 and 12 months respectively, irrespective of antigen specificity (figure 3B).

**Figure 4.**
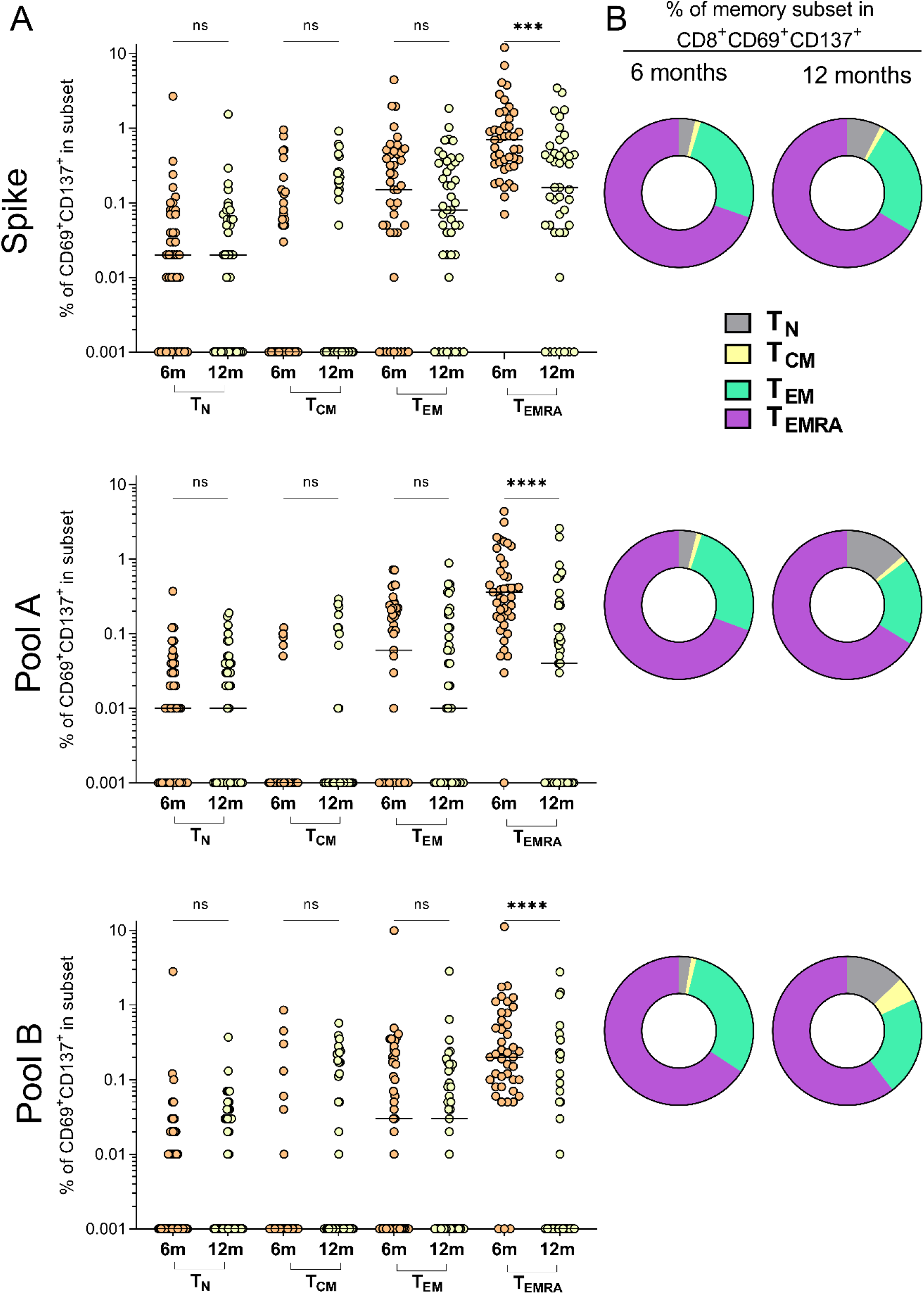
Memory immunophenotyping and quantification of SARS-CoV-2-specific CD8^+^ T cells in mild-COVID-19 convalescents at 6 and 12 months after PCR positive test. **(A)** Memory immunophenotype based on expression of surface CCR7 and CD45RA of SARS-CoV-2 Spike-, whole proteome pools A- and B-specific CD8^+^ T cells detected in Figure 2B. Immune phenotypes were defined as in Figure 3. Frequencies are indicated as percentage of total SARS-CoV-2 antigen-specific CD8^+^ T cells within the total pool of immune cells with same phenotype in the patient’s PBMCs.**(B)** Doughnut charts indicating the proportion (%) of each immune phenotype in (A) of the total of SARS-CoV-2-specific CD8^+^ T cells for each antigen. In (A) circles represent patient individual values. Averages are denoted by a horizontal line and statistically significant differences between time points indicated by asterisks. ^***^ and ^****^ denote *P* values < 0.05 and 0.01, respectively. ns = not significant.

Analysis of the SARS-CoV-2-specific memory CD8^+^ T cells revealed important differences, compared to CD4^+^ T cells. Predominant CD8^+^ T_EMRA_cells specific to SARS-CoV-2 suffered a pronounced reduction regardless of antigen specificity. The frequency of Spike-specific CD8^+^T_EMRA_ decreased from 0.66-1.96 to 0.23-0.70 (95% CI, p<0.001), as well as for the for SARS-CoV-2 whole proteome, (from 0.38-0.92 to 0.07-0.38, p<0.001, for pool A) (from 0.12-1.18, to 0.02-0.34, p<0.001, for pool B) (figure 4A). The importance of T_EMRA_ cells within the CD8^+^ T pool in COVID-19 convalescents was further highlighted as they were the predominant proportion of CD8^+^CD69^+^CD137^+^ cells at 6 months (>65%) and at 12 months (>60%), irrespective of antigen specificity (figure 4B).

The presence of significant frequencies of SARS-CoV-2 antigen-specific CD4^+^ T_EM_ and CD8^+^ T_EMRA_ at 12 months, has important implications, particularly in the absence of virus re-exposure, given that in principle these cells have a limited life-span. Importantly, a significant decrease is these populations from 6 to 12 months suggests that in the absence of restimulation by either infection or vaccination, these populations will eventually not be detectable in circulation, or plateau at low frequencies for extended periods of time. In future, the renewal mechanisms of both of these cell populations is an important question that needs to be addressed.

### Functionality of Spike-specific T cells in ‘high-responder’ convalescents

A direct indicator of of T cell effector function is their ability to produce immunomodulatory cytokines upon antigen stimulation. Since Spike was the immunodominant SARS-CoV-2 antigen according to our AIM results (figure 2), we selected COVID-19 convalescents with Spike-specific CD4^+^ and CD8^+^ T cell responses well above healthy control levels (mean plus 3^*^standard deviations) (figure 5A), hereafter referred to as ‘**high responders’**. We performed a Spike-specific CD4^+^ and CD8^+^ T cell intracellular cytokine staining (ICS) assay measuring the pleiotropic signalling cytokines interferon gamma (INFγ) and tumor necrosis factor alpha (TNFα), as well as the cytotoxic enzymes, granzyme B (GZMB) and perforin (PRF1), and T cell-activation factor, interleukin-2 (IL-2). Additionally, we evaluated the frequency and functionality of Spike-specific circulating T follicular helper cells (cT_FH_) given their importance for immune memory in COVID-19 convalescents^22,52^ (see figures S5 and S6 for ICS/cT_FH_ gating strategy).

**Figure 5.**
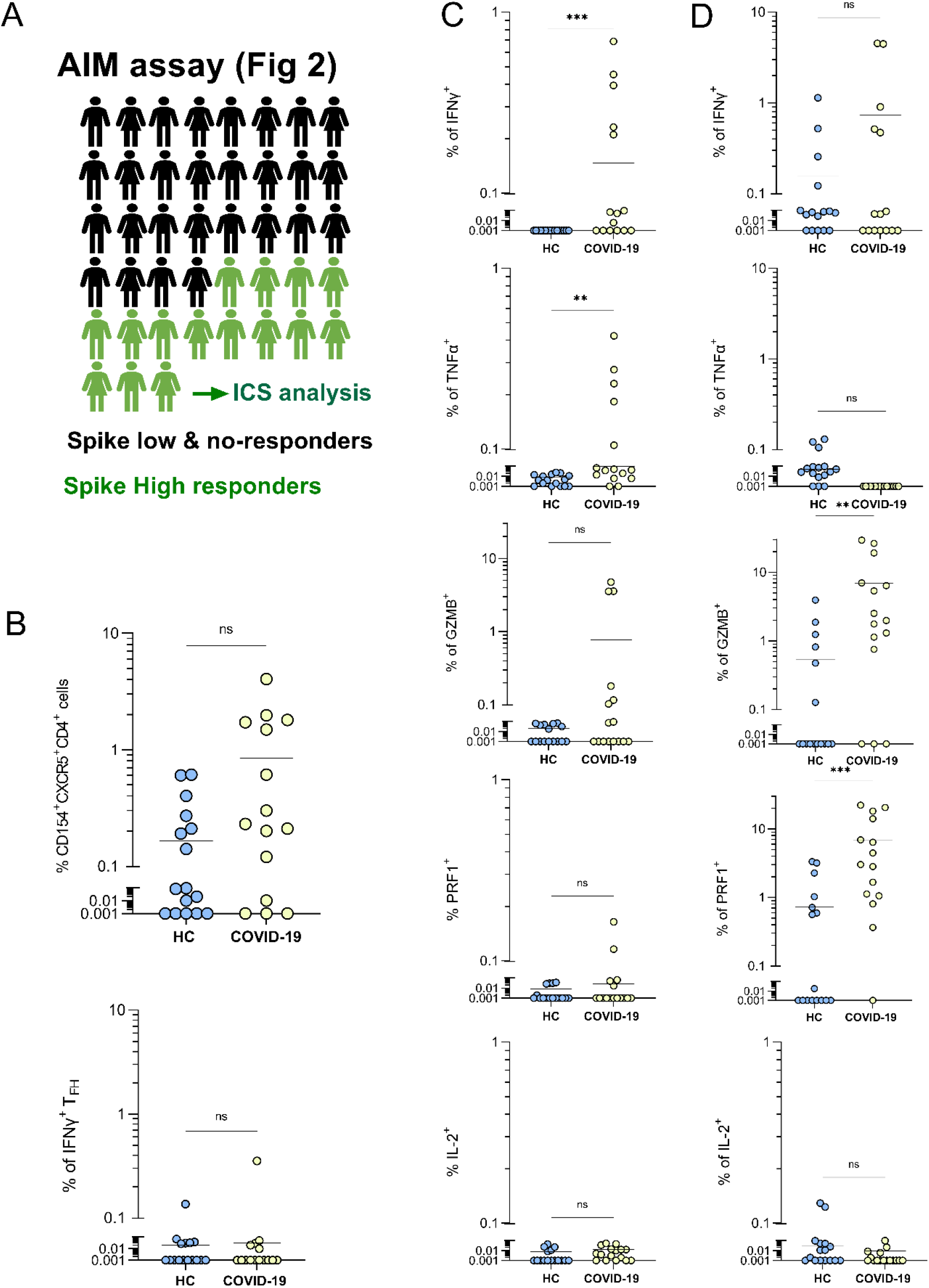
Cytokine expression and circulating T follicular helper cell frequencies in convalescents with strong T cell immune responses to SARS-CoV-2 Spike protein. (A) Fifteen Spike “high responder” convalescents were selected based on the criterion of >healthy control average +3SD of Spike-specific CD4^+^ and CD8^+^ T cell frequencies in the AIM assay (Figure 2) and further analysed for Spike-specific cT_FH_ and ICS analysis using the whole Spike peptide pool.**(B)** Frequency of circulating Spike-specific T follicular helper cells (CD4^+^CXCR5^+^iCD154^+^) in healthy controls (n=15) and selected high responder convalescents (n=15) and % of IFNγ^+^ cells.**(C)** Percentage of cytokine positive expression in CD3^+^CD4^+^iCD154^+^ T cells in same individuals as (B).**(D)** Percentage of cytokine positive expression in CD3^+^CD8^+^ T cells in same individuals as (B). In B, C and D circles represent healthy or patient individual values. Averages are denoted by a horizontal line and statistically significant differences between time points indicated by asterisks. ^**^ and ^***^ denote *P* values < 0.01 and 0.001, respectively. ns = not significant.

At 12 months post-infection, the frequency (%) of Spike-specific cT_FH_ cells in COVID-19 convalescents was not significantly different (95% CI, 1.48-0.21) to that of healthy controls (0.27-0.05) (figure 5B). The frequency of IFNγ^+^ (figure 5B),TNFα, GZMB, PRF1 or IL-2 (figure S7) positive cT_FH_ cells also did not differ from that of healthy controls. This is, to our knowledge, the first report of cytokine expression data in cT_FH_ in COVID-19 convalescents at 12 months post infection.

The reactivity of CD4^+^ T cells in high-responder convalescents to the whole Spike protein was dominated by a higher percentage of CD4^+^ T cells expressing IFNγ (95% CI, 0.26-0.02, p<0.001) and TNFα (0.16-0.02, p<0.01) (figure 5C). The percentage of CD4^+^ T cells expressing GZMB, PRF1 and IL-2 were not significantly higher than in healthy controls (figure 5C). In agreement with their known cytotoxic function, Spike-specific CD8^+^ T cells in COVID-19 convalescents exhibited high frequencies of GZMB (95% CI, 12.30-1.43, p<0.01) and PRF1 (11.27-2.56, p<0.001) while IFNγ, TNFα or IL-2 expressing cells were not significantly different (figure 5D). Taken together, our AIM and ICS data clearly show a durable, multifunctional response of circulating helper and cytotoxic T cells 12 months post-COVID-19 infection. Importantly, we show that cT_FH_ frequencies and their effector function were diminished.

### T cell specificity to VoC Spike mutated sequences in mild-COVID-19 ‘high responders’

Recent studies suggest that T cell specificity to whole Spike antigen and other SARS-CoV-2 proteins is relatively unaffected by the changes in Spike amino acid sequence characteristic of current VoC.^25^ However, these studies have either used peptide pools covering both mutated and non-mutated regions of individual antigens (Spike and non-Spike),^25^ in which mutated peptides represent a very small proportion of the overall pool. Hence any differences in T cell specificities and functionality against the mutations in the Spike antigen of VoC were potentially masked. Others, used mutation-specific Spike peptides, but only for B.1.1.7 and B.1.351 VoC.^26^ Here, we have tested T cell specificity and functionality to the Spike protein of all four VoC with peptide pools covering corresponding mutated regions by intracellular cytokine staining in ‘high-responder’ COVID19 convalescents (n=15) matching reagents and analysis strategies (figures S7 and S8) from previous studies to allow for relevant comparisons.^25,26^

For the context of this experiment and its results, it is important to note that our cohort was infected in early 2020 in Australia, where SARS-CoV-2 with unmutated Spike amino acid sequences was prevalent (figure 6A). In order to clearly identify any changes in functionality of T cell immunity to the Spike antigen present in the current VoC, we set a 2-fold decrease (50%), as a threshold to compare the T cell reactivity to mutated Spike peptide pools with that to reference (unmutated) Spike pools (denoted by a red dotted line in figures 6B and 6C). We show that CD4^+^ T cell cytokine responses were significantly more affected by mutations in VoC Spike antigens, compared to CD8^+^ T cell cytokines responses (figures 6B and 6C). There were 56 instances where a convalescent showed >2-fold reduction for one of the cytokines for one VoC for CD4^+^ T cells compared to 14 for CD8^+^ T cells. Mutations present in the Spike antigen of B.1351 variant induced the highest number of >2-fold reductions in CD4^+^ T cell cytokine/VoC responses in the ‘high responder’ cohort with 23 (41%) of the all (56) of such reductions followed by B.1.617.2 (15, 26%) (figure 6B). These reductions occurred at higher frequency for IFNγ responses in CD4^+^ T cells (16) compared to TNFα, GZMB, PRF1 or IL-2 (12, 10, 11 and 7, respectively) (figure 6B). Only INFγ production was significantly affected in CD8^+^ T cells, with (9 reductions) and no obvious differences in terms of VoC driving these differences (figure 6C).

**Figure 6.**
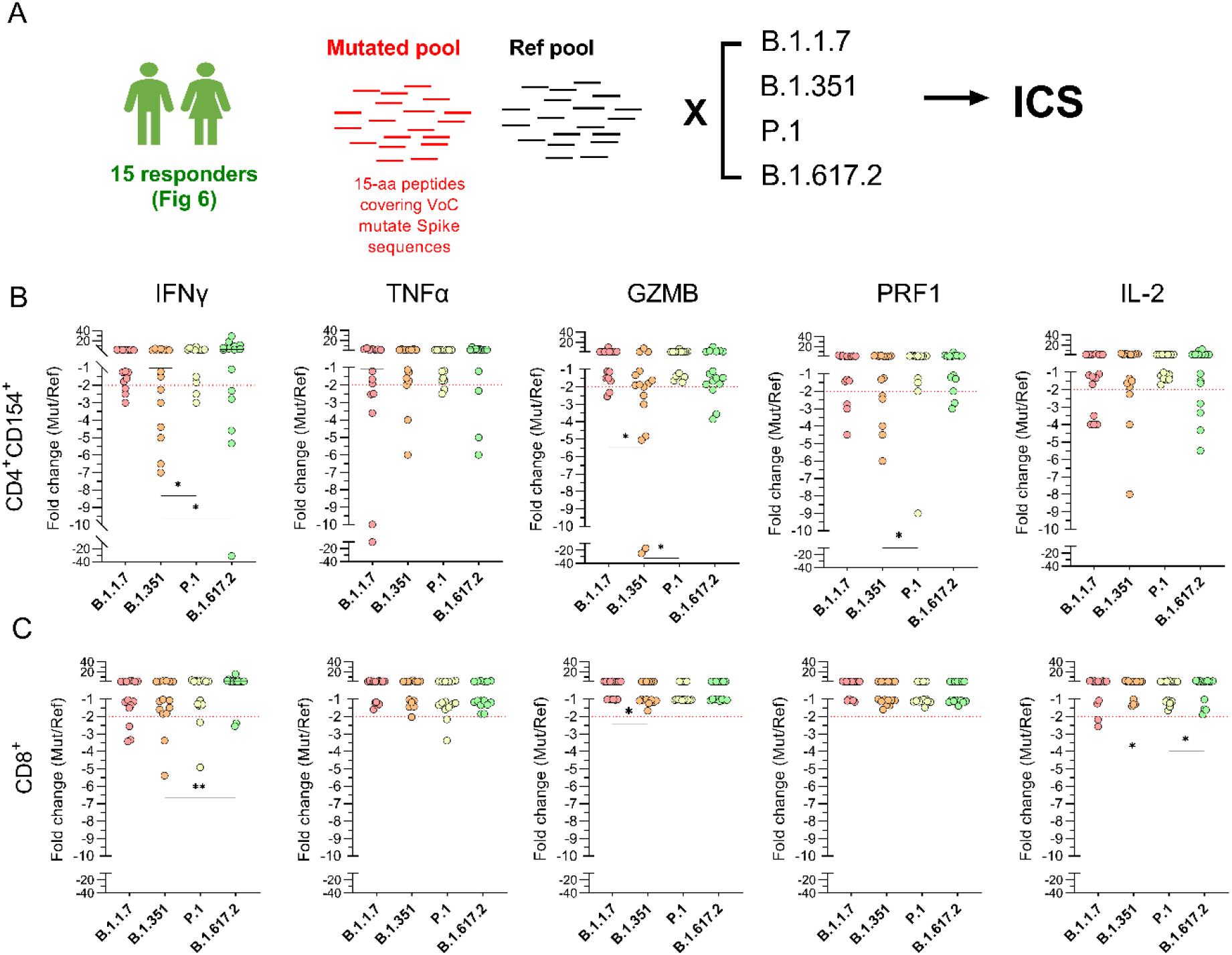
Specific T cell responses in responder convalescents to mutated Spike amino acid sequences in VOCs. **(A)** PBMCs from spike responders (n=15, same as in figure 5) were stimulated with peptides covering mutated Spike regions in VoCs and cytokine expression was measured by intracellular cytokine staining (ICS). **(B)** Fold-change cytokine expression in CD3^+^CD4^+^iCD154^+^ T cells. Negative Folds denote decrease in cytokine expression when stimulated with mutated petide pools. Red dotted line indicates a 2-fold decrease in cytokine expression with corresponding mutated peptide pool. **(C)** Fold-change cytokine expression in CD3^+^CD8^+^ T cells. Negative Folds denote decrease in cytokine expression when stimulated with mutated peptide pools. Red dotted line indicates a 2-fold decrease in cytokine expression with corresponding mutated peptide pool. ^*^ and ^**^ denote *P* values < 0.05 and 0.01, respectively. Non statistically significant differences were not indicated for clarity.

Strikingly, when the same data was analysed looking at individual COVID-19 convalescent participants, 86% of ‘high responders’ exhibited at least one deficient T cell cytokine response (>2-fold) for one of the VoC (table S2). In these convalescents, 48% of all cytokine T cell deficient responses occurred in 4 out of 15 convalescents, clearly indicating differences in the vulnerability to VoC of different convalescents even within a group of ‘high responders’ (table S2). To our knowledge, this is the first time that a loss of Spike-specific T cell functionality against VoC, despite durable (12 months) maintainance in overall frequency, has been reported in COVID-19 convalescents. These data suggest that suboptimal Spike-specific T helper cell functional responses are more likely to occur in convalescents (infected with Wuhan-like variants) who encounter B.1.351 or B.1.617.2, rather than other VoC. This is similar to what happens with humoral antibody responses in COVID-19 convalescents and vaccinees,^12, 18^ and is further supported by our live-virus neutralisation data (figure 1E).

## Discussion

The evidence from large observational studies in healthcare workers and the general population suggests that SARS-CoV-2 immunity post infection confers a level of protection against COVID-19.^53-55^ One caveat of these studies is that high rates of transmission of SARS-CoV-2 in the communities where the studies were conducted (*e*.*g*. Italy, UK or USA) increased the likelihood of virus re-exposure in that cohort, precluding the ability to draw firm conclusions on the duration and protective effects of primary SARS-CoV-2 immunity.

Controlled, in-depth studies in convalescents have also been conducted, but vaccination and almost ubiquitous high SARS-CoV-2 infection rates have reduced the opportunities to recruit convalescents long after COVID-19 disease, and importantly, in the absence of re-infection.

Conversely, vaccination programs allow for more controlled studies and deeper immune analysis and facilitate conducting experimental procedures to test immune fitness against VoC. However, to date, the majority of vaccinees have received Spike-based vaccines, which are not well-suited as a proxy for what SARS-CoV-2 immunity may look like in the long-term, or its adaptability to VoC. Here, we have taken advantage of the relatively unique situation in South Australia, where local transmission of SARS-CoV-2 was eliminated early on in the pandemic in 2020, enabling us to conduct a 12-month longitudinal study of SARS-CoV-2 immunity in mild-COVID-19 convalescents and test their immune fitness against the current VoC.

Total RBD IgG titers and other circulating Ig isotypes against SARS-CoV-2 Spike and RBD antigens decrease over the months following COVID-19 disease, with the strongest decline in the first 1-3 months.^22,36,56^ A concordant trend was also observed in our cohort between 6 and 12 months. A recent study in Wuhan (China), where RBD-specific IgG was longitudinally assessed in convalescents up to 12 months post-COVID-19 in a community with absence of SARS-CoV-2 local transmission, analogous to that of South Australia, showed a similar trend between 6 and 12 months.^9^ It is worth noting that the genome sequence of the virus variants causing COVID-19 in each of the studies are closely related and in the absence of VoC. Despite declining circulating antibody titers, the majority of convalescents in our cohort (>90%) had significantly higher levels of Spike- and RBD-specific total IgG compared to the healthy controls at 12 months post-COVID-19. Concomitantly, circulating RBD-specific memory B cells were present in 88.9% of convalescents, indicating that antibody responses could extend further into the future.

The presence of specific antibodies alone does not necessarily predict protection against disease. Mathematical modelling of clinical data indicates that a minimum specific Ig titer is needed for protection against COVID-19 disease.^57^ This is also supported by clear evidence showing that serum virus neutralisation activity, exerted by a portion, but not all antibodies, is the best predictor of protection.^48,55,57,58^ Both live-virus and pseudovirus serum neutralisation assays indicated that sera from 51% and 65%, respectively, of COVID19 convalescents can efficiently neutralise SARS-CoV-2 bearing the Spike protein homologous to one which caused the original infection, 12 months post-infection. At 6-8 months post-COVID-19, the percentage of convalescents who retain serum neutralizing activity, estimated with methods similar or identical to ours, varies considerably, from ∼60% to >90%.^8,22,36,43^ It is important to note that these studies included more heterogenous cohorts, which included severe COVID-19 patients. Overall, virus neutralisation measures in our cohort (51-65%) 12 moths post-infection are in line with previous findings.

Recent modelling data predicted the risk of reinfection in COVID19 convalescents to emerging variants of SARS-CoV-2 and showed that reinfection likelihood is high, at median time of 16 months after primary infection, earlier than for other human coronaviruses.^59^ Empirical data is needed to confirm these predictions to better manage the ongoing pandemic.

We demonstrated with live-virus neutralisation data that despite the maintenance of serum antibodies at 12 months post infection in 90% of mild COVID-19 convalescents, they have significantly decreased ability to neutralise the four main VoC. It is striking to see that serum neutralising activity against B.1.351 is practically non-existent, and for both P.1 and the pervasive variant B.1.617.2, the drop in the number of patients who retain serum neutralisation activity is >65%. For B.1.1.7, the first VoC (chronologically), and perhaps less relevant at the present time, the reduction in neutralisation titers was less pronounced. The relative differences at 12 months post-infection between the VoC correlated well with other COVID-19 convalescent and vaccination studies.^13,14,16,18,57,60-62^ Our measurements at 12 months post-COVID-19 and in the absence of re-infection, reveal that although circulating SARS-CoV-2 antibody responses persist up to 12 months, the antigenic drift of SARS-CoV-2 in VoC outperforms humoral immune responses in mild-COVID-19 convalescents.

As such, our findings stress the necessity to reinforce the immune system to overcome these insufficiencies in COVID-19 convalescents via vaccination.

Importantly, we report that SARS-CoV-2-specific circulating CD4^+^ and CD8^+^ T cell frequencies are maintained in a significant proportion of convalescents between 6 and 12 months post-COVID-19 and at levels reported for earlier time points,^22,23^ indicating that these T cell populations are stable. While >50% of SARS-CoV-2-specific CD4^+^ and CD8^+^ T cells were directed at the Spike protein, a significant proportion of T helper and cytotoxic T cells were activated by non-Spike antigen epitopes.

Collectively, memory compartment analysis of circulating SARS-CoV-2-specific T cells strongly suggests that in mild-COVID-19 convalescents, CD4^+^ T_EM_ and CD8^+^ T_EMRA_ have significant roles in maintaining immunity as far as 12 months after infection. Interestingly, the maintenance and large proportion of CD8^+^ T_EMRA_ bears resemblance to the kinetics of equivalent cells in long-term chronic infections by human CMV.^63^ A significant decay of Spike-specific CD4^+^ T_CM_ cells was observed, however this was not reflected in their CD8^+^ T cell counterparts, perhaps indicating that in the long term, cytotoxic immune function could be sustained more efficiently thanks to cell self-renewal and proliferation.^51^ Importantly, in our cohort of ‘high responders’, T cells showed a strong capacity to produce a range of cytokines, with strong IFNγ and TNFα responses in CD4^+^ T cells and GZMB and PRF1 in CD8^+^ T cells, in concordance with their respective helper and cytotoxic function profiles.

At 12 months post-infection we did not detect significant frequencies of circulating Spike-specific cT_FH_ cells, even in ‘high-responder’ convalescents, which is in agreement with studies that showed their decline in earlier months after infection^22^ and vaccination.^64^

The current understanding of SARS-CoV-2 infection-induced T cell responses with respect to their ability to adapt to VoC with Spike antigen mutations, is that contrary to antibody responses, T cells significantly maintain reactivity to such mutated antigens.^25,26^ In contrast, we show, with a functional cytokine T cell assay, that changes in the Spike amino acid sequence corresponding to VoC, led to functional vulnerabilities, where even ‘high responder’ COVID-19 convalescents exhibited reduced cytokine production in response to mutated Spike antigen epitopes. Importantly, we found that a subset of ‘high responder convalescents’ accumulated most of these functional deficiencies in their CD4^+^ T cell compartment, rather than cytotoxic CD8^+^ T cells, with such deficiencies more pronounced against B.1.351 Spike epitopes.

The ability of viruses to escape virus-specific T cell responses is known.^65,66^ In fact, viruses such as influenza can escape T cell responses after acquiring single amino acid mutations.^67^ Hepatitis C virus is also capable of escaping T cell responses with minimal variations to antigen amino acid sequences.^68^ Our findings are concerning, since additional mutations leading to the appearance of new SARS-CoV-2 variants could potentially increase the ability of the virus to escape pre-existing Spike-specific T cell responses.

Our results regarding T cell adaptability to VoC are limited in that we have only tested cytokine responses to a single antigen, Spike. Our AIM assay results clearly show that Spike is the main driver of immune responses in convalescents, but immunity to other non-Spike proteins is also important. Variation in direct T cell effector function (cytokine production) to corresponding non-Spike antigenic VoC sequences was not tested and, therefore, the combined SARS-CoV-2 protein effect of VoC mutations on T cell immune function is yet to be fully elucidated. Prior to this study, Tarke *et al* conducted multiantigenic studies (including, Spike, N, M, and a range of ORFs and NSPs) to test T cell responses to VoC (excluding the prevailing B.1.617.2 variant), concluding that T cell reactivity to ancestral Wuhan antigen epitopes and virus variants was not significantly different. In their experimental design, they used an AIM assay-based approach with large peptide pools, which included mutated and non-mutated epitopes. While this approach bears more resemblance to what might happen during a SARS-CoV-2 infection, it potentially masks subtle differences in T cell reactivity and the identification of individuals whose T cell responses are more severely affected by antigenic changes in the Spike antigen of VoC.^25^ Also, while the AIM assay is a powerful method to detect and quatify T cell antigen specificity it does not measure function directly, compared to ICS.

On the other hand, while testing Spike alone may be a limitation when assessing convalescent responses, our results have important implications in assessing immunity elicited by most of the current vaccines administered in developed countries, which are based, exclusively, on the Wuhan-Spike protein (e.g., vaccines produced by AstraZeneca, Pfizer-BioNTech, Janssen or Moderna). If the deficiencies reported here have an equivalent in uninfected vaccinees, then, functional T cell responses could be compromised in these individuals. Future studies must address this question.

In summary, our findings reveal that, despite the durability and maintenance of serum antibodies, circulating memory B cells and T cell responses at 12 months after original infection, COVID-19 convalescents have pronounced deficiencies in functional Spike-specific T cell responses and the ability to neutralise the current VoC. As such, mild-COVID-19 convalescents are vulnerable to infection with circulating and newly emerging SARS-CoV-2 variants 12 months after recovery. The results obtained from our functional evaluation of post-COVID-19 immunity are in line with current large observational studies, that demonstrate the added protective effect of COVID-19 vaccines in previously infected individuals.^69,70^ Thus our study highlights the importance and necessity to vaccinate COVID19 convalescents against subsequent SARS-CoV-2 infections.

Finally, it is important to consider our findings in the context of the existing worldwide inequalities in vaccine distribution, which have kept many developing countries from Africa, South East and South Central Asia and Central America at vaccination rates <20% so far.^71^ These regions are, therefore, more vulnerable to outbreaks of emerging SARS-CoV-2 variants with the ability to evade pre-existing immune responses in unvaccinated convalescents.^72, 73^

## Data Availability

All data produced in the present study are available upon reasonable request to the authors

## Author contributions

Writing original manuscript draft: PGV, BGB and MGM. Editing manuscript: SCB, MRB, CKL, EJG, GM and DJL. Design of experiments: PGV, CMH, MGM, AS, DW, SGT, BGB, SCB and RAB. Performed experiments: PGV, CMH, MGM, AELY, HB, ZAM, ZAD, AA, AOS, AA and DA. Data analysis: PGV, CMH, AELY, BGB, RAB. Patient sample collection: CMH, JG, CF, SO, EMM, BAJR, DS, and CKL. All authors reviewed, discussed, and agreed with manuscript.

## Acknowledgements

This work was funded by project grants from The Hospital Research Foundation and Women’s and Children’s Hospital Foundation, Adelaide, Australia. MGM is THRF Early Career Fellow. BGB is THRF Mid-Career Fellow. This work has been supported by NIH contract 75N9301900065 (A.S, D.W). We would like to thank Ms. Suzanne Edwards, University of Adelaide, for statistical advice and analysis.

## Supplementary Information

**Table S1.**
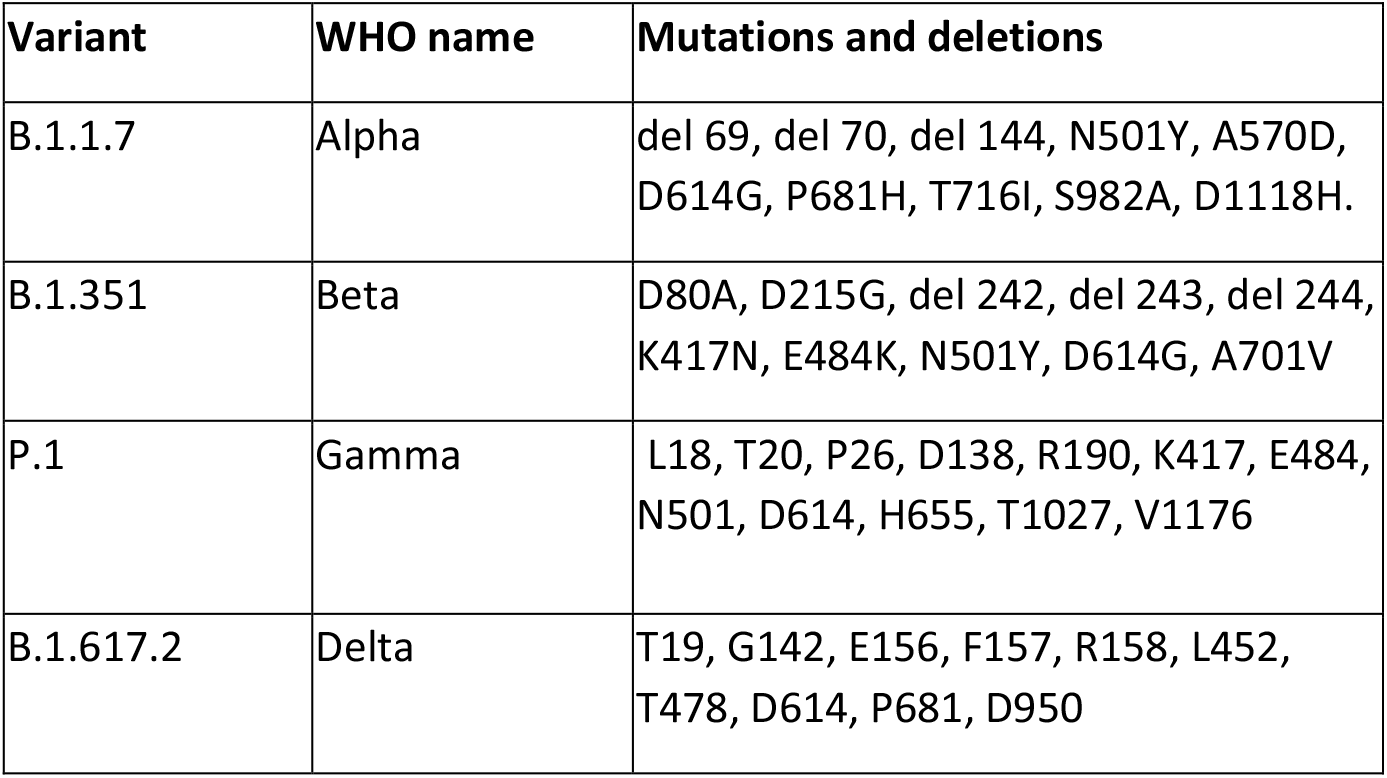
PepTivator® peptide pool mutation and deletions.

**Table S2.** No. of times each high-responder convalescent exhibited cytokine T cell frequencies reduced by >2-fold when stimulated with any of the mutated VoC Spike peptide pools in figure 6.

| ID | CD4+<br>cytokine | CD8+<br>cytokine | total |
| --- | --- | --- | --- |
| 15-006 | 5 | 0 | 5 |
| 15-008 | 11 | 0 | 11 |
| 15-012 | 6 | 3 | 9 |
| 15-013 | 2 | 0 | 2 |
| 15-016 | 4 | 1 | 5 |
| 15-020 | 5 | 1 | 6 |
| 15-030 | 2 | 2 | 4 |
| 15-046 | 4 | 0 | 4 |
| 15-054 | 4 | 2 | 6 |
| 15-056 | 0 | 0 | 0 |
| 15-058 | 5 | 2 | 7 |
| 15-059 | 4 | 3 | 7 |
| 15-062 | 0 | 0 | 0 |
| 15-065 | 1 | 0 | 1 |
| 15-070 | 3 | 0 | 3 |

**Figure S1.**
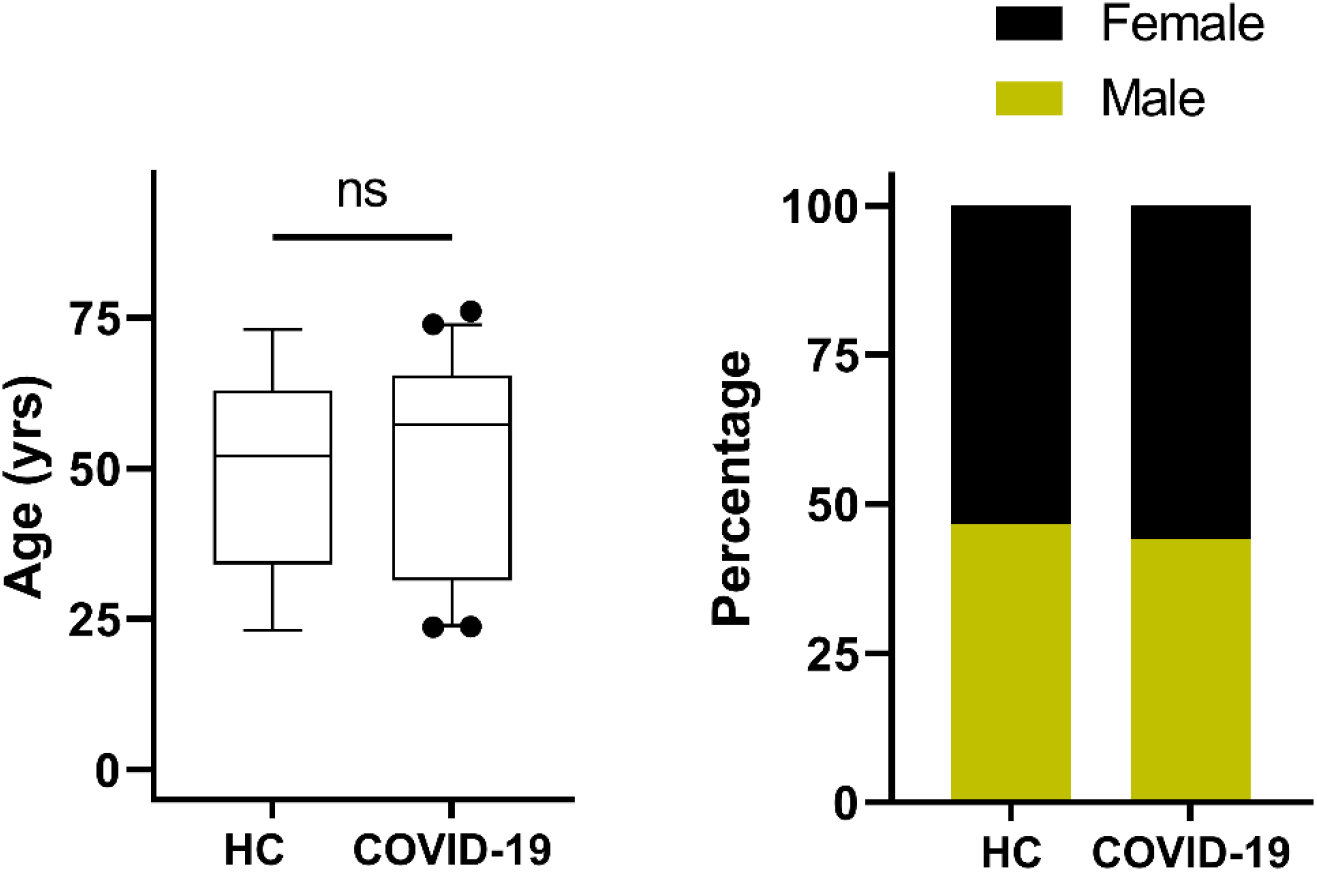
Age and gender distribution of COVID-19 convalescents and healthy control subjects recruited for this study.

**Figure S2.**
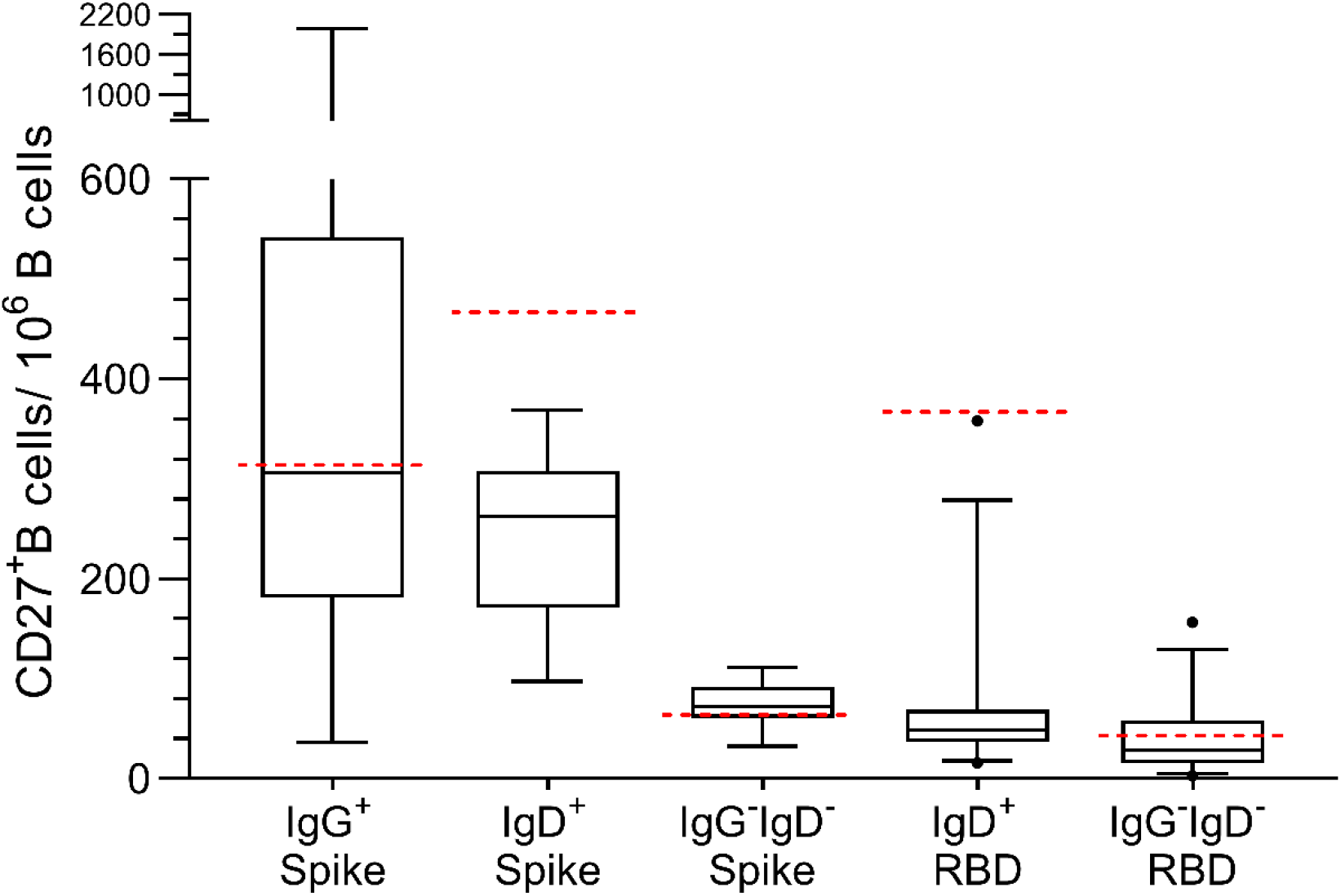
Spike and RBD-specific memory B cells in COVID-19 convalescents. The number of Spike and RBD-specific IgG^+^, IgD^+^ or IgG^-^IgD^+^ memory (CD27^+^) B cells per million B cells in COVID-19 convalescents at 12 months post disease onset was evaluated by tetramer staining. Data represented as 5-95 percentile box plots and standard deviation error bars. Staining average + 2^*^ standard deviation values obtained in healthy (uninfected) controls are denoted by dashed red lines in each case.

**Figure S3.**
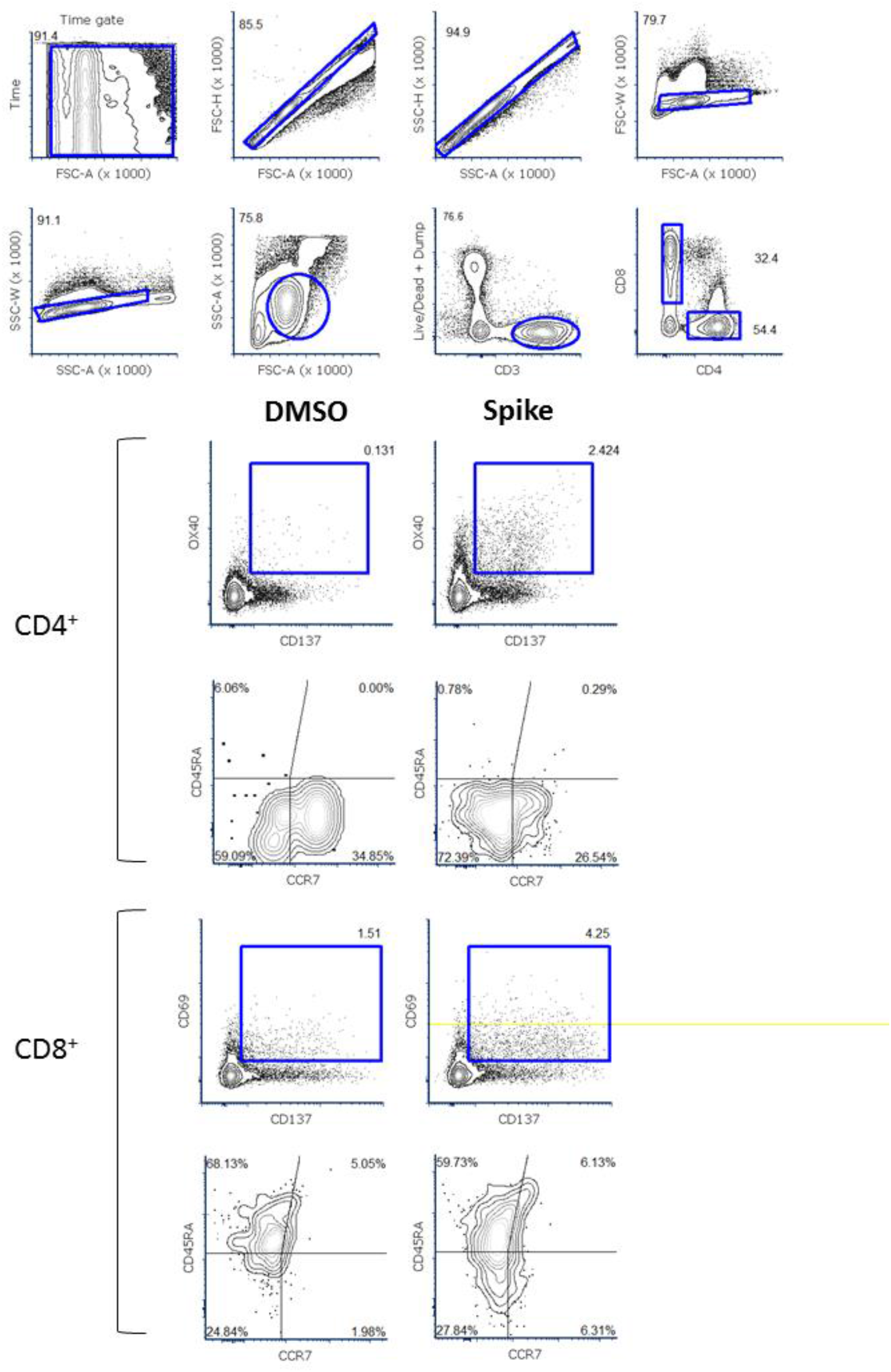
AIM assay gating strategy. Live, CD14^-^CD19^-^ CD3^+^ singlets were gated and further classified as either CD4^+^ or CD8^+^ T cells using gating strategy on two top row panels. Antigen specific CD4^+^ and CD8^+^ T cells were identified based on double expression of surface CD137 and OX40 or CD137 and CD69, respectively (lower panels). Finally, memory phenotype of antigen-specific cells was further determined according to expression of surface CCR7 and CD45RO.

**Figure S4.**
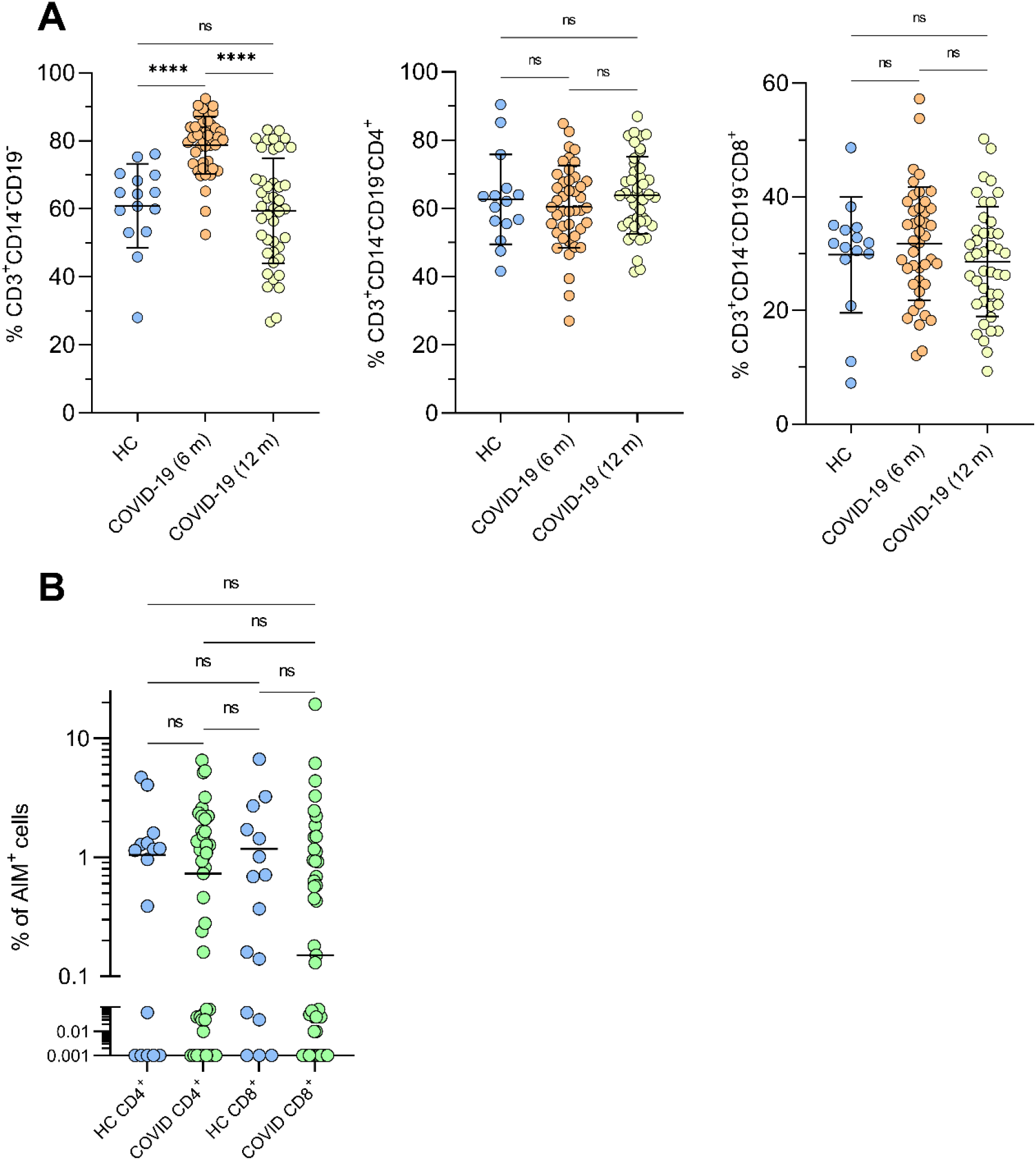
T lymphocytes and CMV-specific CD4+ and CD8+ cells in study participants. (A) Left to right, percentage of CD3^+^, CD3^+^CD4^+^ and CD3^+^CD8^+^ T cells in COVID-19 convalescents at 6 and 12 months after disease onset (n=43) and healthy controls (n=15) analysed in the AIM assay. (B) CD4^+^ and CD8^+^ CMV-specific T cells in healthy controls and COVID-19 convalescents at 12 months after SARS-CoV-2 infection. ^****^ and ns denote statistically significantly differences at p <0.0001 and not significant, respectively.

**Figure S5.**
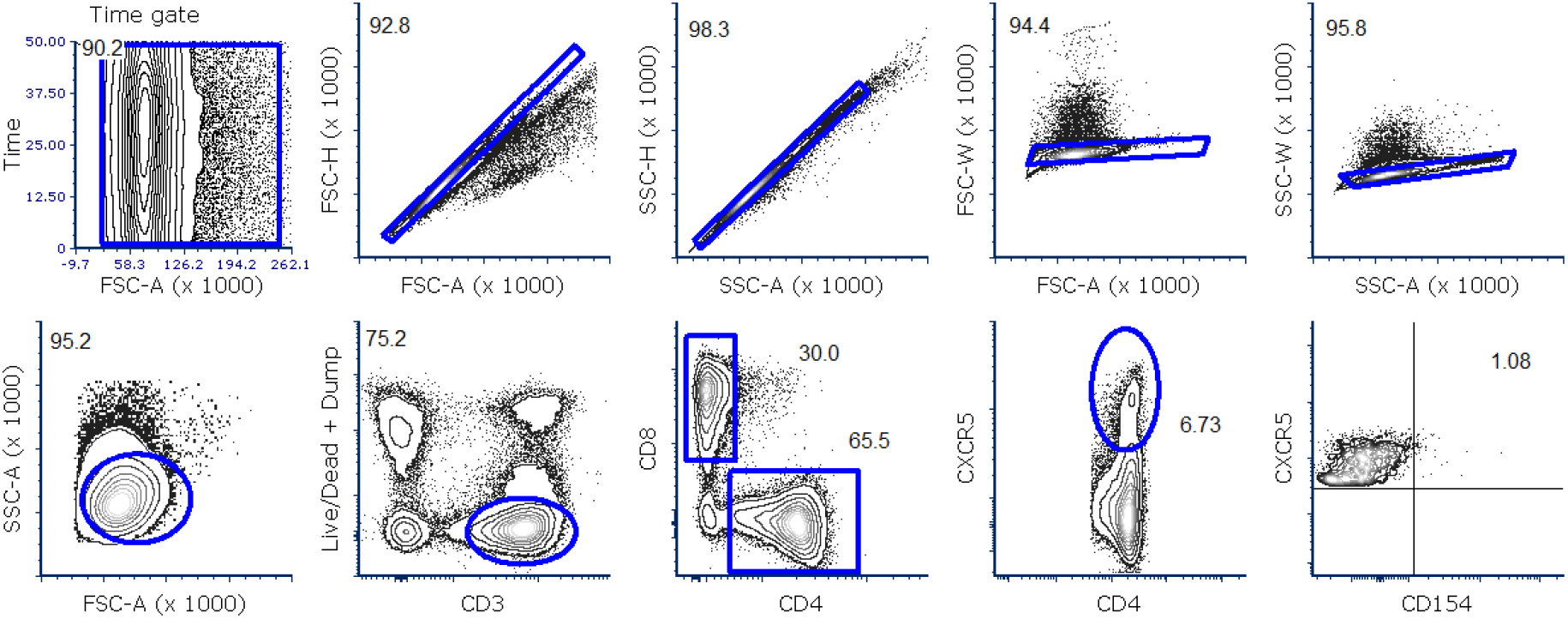
Flow cytometry data gating strategy for whole Spike (Wuhan)-specific intracellular cytokine (ICS) assay and activated T Follicular Helpers. Representative strategy to define CD4, CD8 and T Follicular helpers (CD4+ CXCR5+) for the intracellular cytokine assay and activation induced CD154 (CD40L) expression. CD154 was measured for activation of T Follicular helpers.

**Figure S6.**
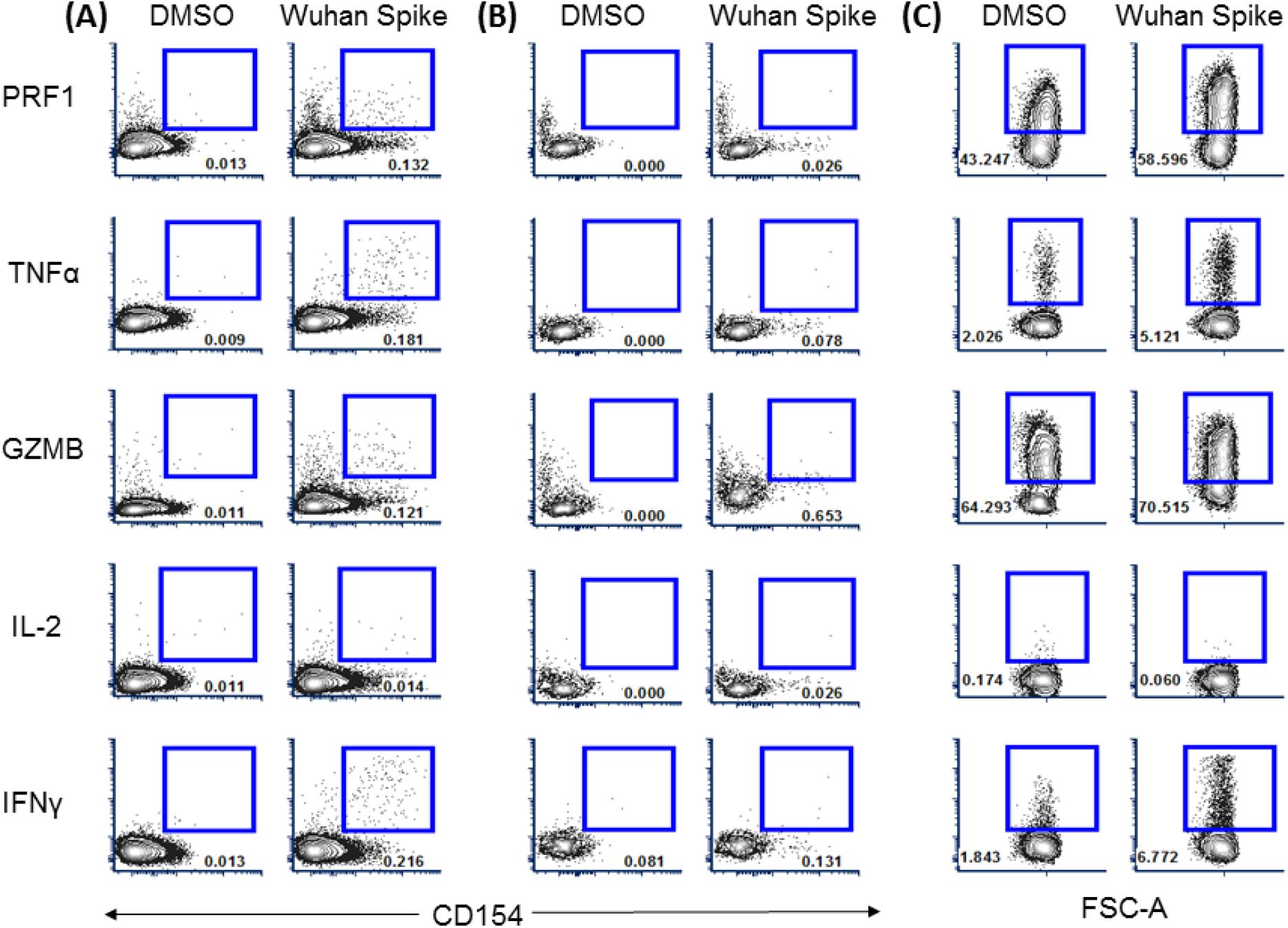
Whole (Wuhan) Spike-specific ICS assay gating strategy. Flow cytometry data gating strategy for Wuhan spike-specific intracellular cytokine (ICS) assay Representative strategy to measure Wuhan-spike-specific (A) CD4, (B) CD4 Tfh and (C) CD8 T cells via percentage of intracellular cytokines: perforin (PRF1), Tumour Necrosis Factor alpha (TNFα), Granzyme B (GZMB), IL-2 and Interferon gamma (IFN-γ). PBMC were stimulated with Wuhan spike megapool.

**Figure S7.**
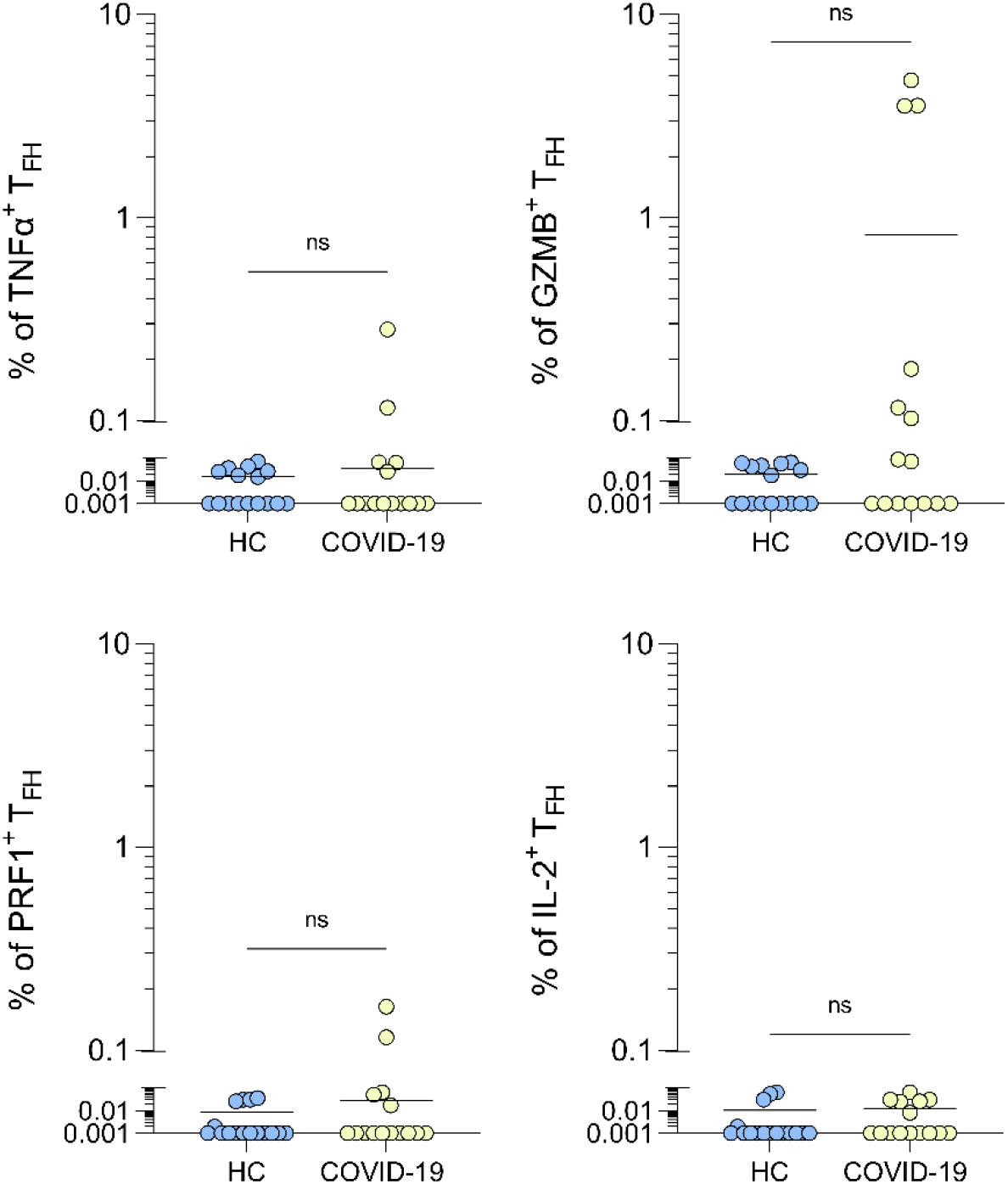
Cytokine expression in Spike-specific T follicular helper cells. Intracellular cytokine expression in circulating T follicular helper cells (CXCR5+CD154+CD4+) in healthy controls (n=15) and COVID-19 convalescents at 12 months after infection (n=15). ns denotes not statistically significant.

**Figure S8.**
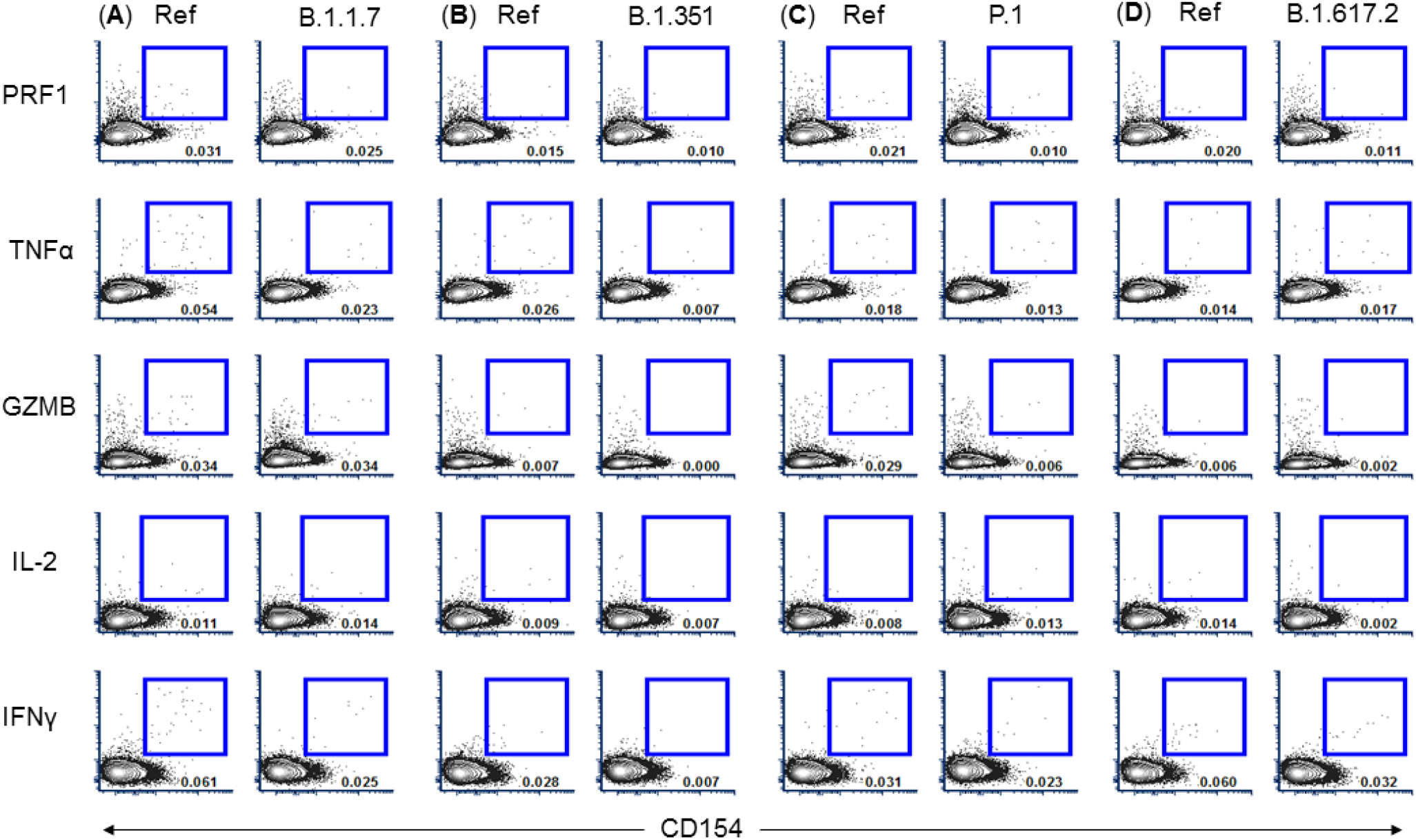
Flow cytometry data gating strategy for Variants of Concern (VoC)-spike-specific intracellular cytokine (ICS) assay for CD4 T cells. Representative strategy to measure Variants of Concern (VoC)-spike-specific CD4 T cells via percentage of intracellular cytokines: perforin (PRF1), Tumour Necrosis Factor alpha (TNFα), Granzyme B (GZMB), IL-2 and Interferon gamma (IFN-γ). PBMC were stimulated with either VoC-specific VoC peptide pools or equivalent peptide pools with corresponding Wuhan aa sequences, that is, reference pool (Ref). (A) Variant B.1.1.7, (B) Variant B.1.3.5.1, (C) Variant P.1 and (D) Variant B.1.617.2.

**Figure S9.**
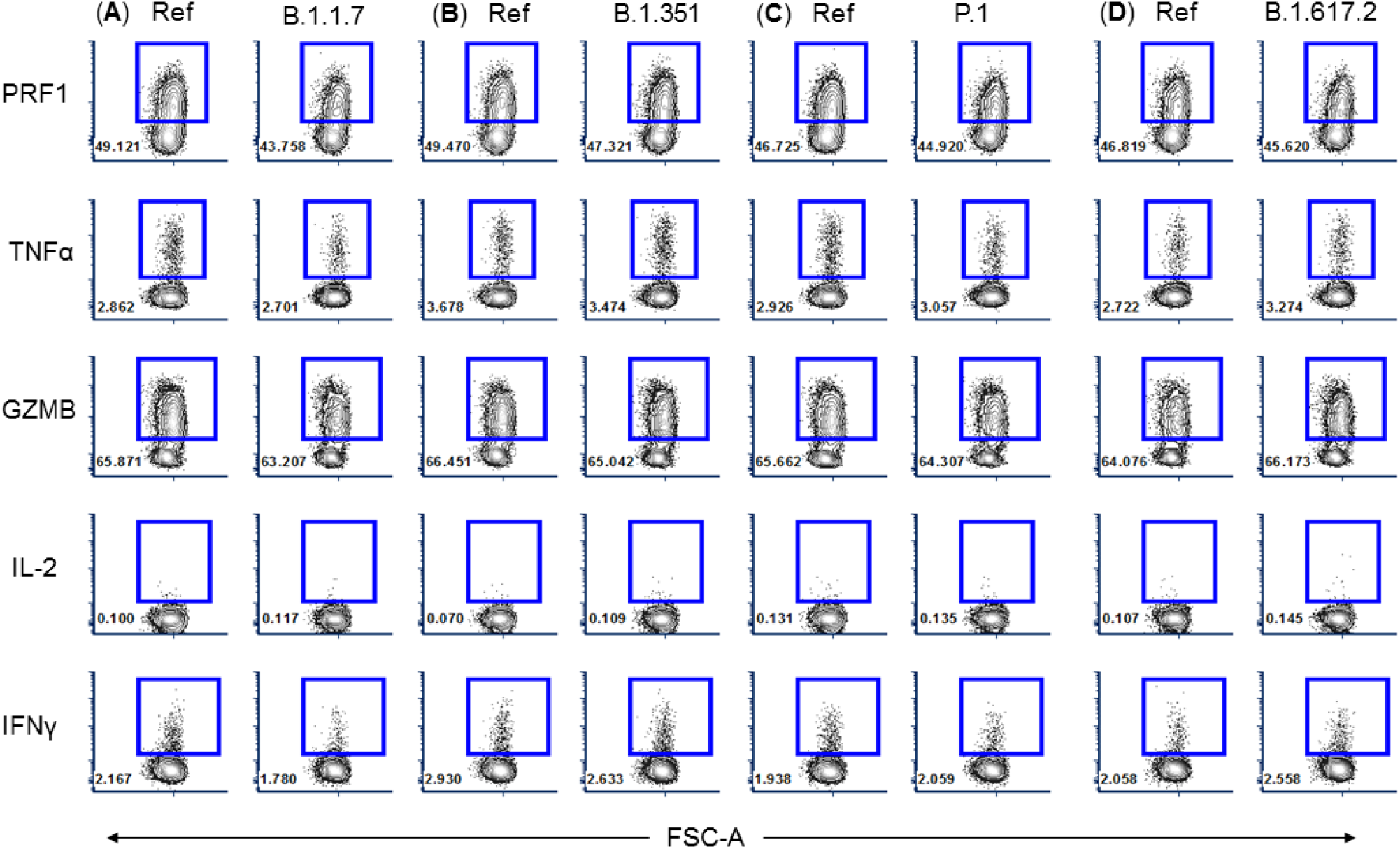
Flow cytometry data gating strategy for Variants of Concern (VoC)-spike-specific intracellular cytokine (ICS) assay for CD8 T cells. Representative strategy to measure Variants of Concern (VoC)-spike-specific CD8 T cells via percentage of intracellular cytokines: perforin (PRF1), Tumour Necrosis Factor alpha (TNFα), Granzyme B (GZMB), IL-2 and Interferon gamma (IFN-γ). PBMC were stimulated with either VoC-specific VoC peptide pools or equivalent peptide pools with corresponding Wuhan aa sequences, that is, reference pool (Ref). (A) Variant B.1.1.7, (B) Variant B.1.3.5.1, (C) Variant P.1 and (D) Variant B.1.617.2.

